# Implementation of a pediatric telemedicine and medication delivery service in a resource-limited setting: A pilot study for clinical safety and feasibility

**DOI:** 10.1101/2022.04.26.22273923

**Authors:** Molly B. Klarman, Katelyn E. Flaherty, Xiaofei Chi, Youseline Cajusma, Lerby Exantus, Jason Friesen, Valery M Beau de Rochars, Chantale Baril, Matthew J. Gurka, Torben K. Becker, Eric J Nelson

**Affiliations:** Departments of Pediatrics and Environmental and Global Health, University of Florida, Gainesville, FL, USA; Department of Emergency Medicine, University of Florida, Gainesville, FL, USA; Department of Health Outcomes and Biomedical Informatics, University of Florida, Gainesville, FL, USA; Université d’État d’Haiti- Faculté de Médecine et de Pharmacie, Port-au-Prince, Haiti; Trek Medics International, Washington, DC, USA; Department of Health Services Research, Management and Policy, University of Florida, Gainesville, FL, USA

**Keywords:** pediatrics, telemedicine, healthcare access, acute, nighttime

## Abstract

**Objective:** Determine the clinical safety and feasibility of implementing a telemedicine and medication delivery service (TMDS) to address gaps in nighttime healthcare access for children in low-resource settings.

**Methods:** We implemented a TMDS in Haiti called ‘MotoMeds’: (i) A parent/guardian of a child ≤10 years contacted the call center (6pm-5am). (ii) A provider used paper clinical decision support tools to triage the case as mild, moderate, or severe. Severe cases were referred to emergency care. For non-severe cases, call center providers gathered clinical findings to generate an assessment and plan. (iii) For households within the delivery zone, a provider and driver were dispatched with medications/fluids; the provider performed a paired in-person exam. For households outside the delivery zone, the family received phone consult alone. All families received a follow-up call at 10-days. Data were analyzed for clinical safety and feasibility.

**Results:** A total of 391 cases were enrolled from September 9^th^, 2019 to January 19^th^, 2021; 89% (347) received a household visit. Most cases were triaged as mild or moderate (92%; 361). Among the severe cases, 83% (20) sought subsequent referred care. The most common complaint was a respiratory problem (63%; 246). At 10-days, 95% (329) of parents reported their child’s condition as “improved” or “recovered”. Ninety-nine percent (344) rated the TMDS as “good” or “great”. The median phone consultation was 20 minutes, time to arrival at the household was 73 minutes and total workflow per case was 114 minutes.

**Conclusion:** The TMDS was a feasible healthcare delivery model with high rates of improved clinical status at 10-days.

**Study registration (clinicaltrials.gov):** NCT03943654

## INTRODUCTION

Stalled progress to improve access to healthcare threatens efforts to meet the United Nations Sustainable Development Goals (SDG). In part, SDG 3.8 seeks to “achieve access to quality essential healthcare services and access to safe, effective, quality and affordable essential medicines for all”;^1^ however, the pace of progress has slowed since 2010 with low-and-middle income countries (LMIC) lagging farthest behind.^2^ Innovative solutions are needed to bypass barriers that are impeding progress towards SDG 3.8.^3 4^

Conventional healthcare models require patients to transit from households to centralized resources. These models are limited by economic, geographic, and logistical barriers, especially at night. Non-conventional approaches that dispatch resources from centralized locations to households represent opportunities to bypass these barriers. Within pediatrics, interventions that prioritize “delivery of services close to home” are associated with a positive improvement in uptake of healthcare.^5^ The WHO/UNICEF action plan for Ending Preventable Child Deaths states that control efforts will be ineffective without innovation within the domains of “delivery strategies, overcoming barriers to interventions, and better ways for implementation”.^6^

Demand for telemedicine increased during the COVID-19 pandemic as it effectuated provider efficiency and increased accessibility for patients.^7-9^ The application of telemedicine in low-resource settings has potential to overcome workforce, distance, and logistical barriers.^10-12^ However, provision of information without medications can be of limited value. Telemedicine coupled with medication delivery likely has the capacity to overcome more access barriers than either service by itself. Use cases of telemedicine and medication delivery services (TMDS) are generally limited to high-income countries^13^, chronic illnesses^14^ and daytime use.^15^ Herein, we describe the implementation of a TMDS called ‘MotoMeds’ for pediatric patients suffering from acute illness at night. MotoMeds was designed to be country agnostic yet customizable to local needs.

The pilot deployment was in Haiti. Haiti was chosen because of need as well as economic, political, and climatic conditions that readily expose weaknesses in healthcare delivery models. MotoMeds is the product of a series of initiatives called the Improving Nighttime Access to Care and Treatment (INACT) studies. INACT-1 was a needs assessment of Haitian households and their providers. Households identified accessibility and affordability as key barriers to healthcare access.^16^ When deciding where to seek care, families cited distance as the primary reason for choosing their provider. Families expressed intentions to seek care from conventional providers (licensed entities) within the larger healthcare network. However, in practice they sought care from a mixture of conventional and non-conventional providers who practice outside of healthcare networks. Non-medical reasons such as financial and logistical challenges (e.g. geographic and nighttime illness) were the primary determinants for deciding when to seek care. These data suggest families are unable to travel far, or pay, to access their preferred healthcare providers, especially at night. Technology solutions emerged as potential mechanisms to bypass these access barriers. In response, MotoMeds was deployed in the same study area as the needs assessment. Our objective was to evaluate the TMDS from clinical safety and feasibility perspectives.

## METHODS

### Ethics Statement

The study protocol was approved by the Comité National de Bioéthique (National Bioethics Committee of Haiti; 1819-51) and the Institutional Review Board (IRB) at University of Florida (IRB201802920).

### Study design

This observational census-based pilot study was designed to monitor clinical safety and assess the feasibility of implementing the TMDS.

### Study population

This study was conducted in Haiti where 23% of the total population and 5% of the rural population have access to quality primary care.^17^ Less than half of the total population lives within 50 km of a tertiary care facility.^18^ The median age is 22 years and life expectancy is 62 years.^19^ The under five mortality rate is 63 per 1,000 live births compared to 38 per 1,000 live births globally.^20^ Acute Respiratory Infection (ARI) and diarrheal disease are common, accounting for over half of hospitalizations and over a third of deaths.^21^ The study was conducted in the commune of Gressier, and consists of a semi-urban and rural agricultural plane with rural mountainous areas and a population of approximately 38,100^22^. The study area had inconsistent 3G cellular coverage. The delivery zone covered approximately 80 sq km; a circle formed from a 5 km radius surrounding the Gressier call center (Figure S1).

### Participant and public involvement

Design of the TMDS and subsequent evaluation was informed by the public through participation in a community based needs assessment on barriers to access healthcare^16^ and through two community stakeholder engagement workshops. Participants passively assisted with recruitment by voluntarily promoting the service via word of mouth.

### Participant recruitment

Recruitment began two weeks prior to launch with radio advertisements, paper flyers, and community engagement at churches, clinics, and markets. Advertising continued throughout the study period. Messaging included the following: the service was available to families with children 10 years or younger experiencing a non-emergent, acute illness during operating hours (6pm-5am), and the TMDS requested a ‘small fee but parents should not let the fee deter them from calling.’

### Participant inclusion criteria, consent process and enrollment estimates

Inclusion criteria were a child 10 years or younger with a medical complaint and an adult parent/guardian caller who was 18 years or older. At the end of the initial call, participants were informed that the TMDS was part of a research study. Written informed consent/assent was performed at the household for those participants receiving a household visit. Participants not receiving a visit were asked to participate in the research study by waiver of documentation of consent over the phone. All callers were informed that their decision to participate would not affect their ability to receive care/ consult. Enrollment was established *a priori* to occur over 12 months (extended to 16 months to meet enrollment) with 571 participants; we estimated 300 participants would reside in the delivery zone and not have danger signs and 271 would reside outside the delivery zone and/or have danger signs. As a pilot, the sample size was based on estimated call-volume. Effect sizes with estimated confidence intervals were calculated based on enrollment and key aspects of the TMDS: rate of identification of dangers signs at the call center and common diagnoses.

### Clinical procedures

In-person WHO Integrated Management of Childhood Illness (IMCI) guidelines were adapted by the study team for use at the TMDS. The resultant tools consisted of first a case report form (CRF) composed of 35 clinical, 4 demographic and 8 operational data points (Supplementary Text 1); clinical decision-support was embedded into the CRF. The second tool was a set of clinical guidelines outlining danger signs, clinical assessments, treatment plans and follow-up recommendations (Supplemental Text 2). These guidelines were written to be used within the scope of this pilot only. The guidelines focused on 6 six common childhood illnesses based on the WHO Global Burden of Disease studies on pediatric morbidity and mortality globally^23^: Fever, ‘Respiratory problem/Cough’, Dehydration/’Vomit’/Diarrhea, ‘Ear pain’, ‘Skin problem’, and ‘Pain with urination’ (Urinary tract infection). Additional complaints were grouped as ‘Other complaints’. Guidelines incorporated different workflows depending on nutrition/malnutrition status. The third tool was a medication formulary of oral and topical medications and fluids (Supplemental Text 3).

All tools were paper-based and adapted to real world use over the course of the study depending on medication availability, changes in national/WHO guidelines and shifts in disease burden (e.g. SARS-CoV-2). The CRF was translated into Haitian Creole and the clinical guidelines and formulary were translated into French. The scope of these tools was not intended to be exhaustive but rather focused on the unique setting of a TMDS. Definitive WHO pediatric reference materials were provided at the call center. If a patient’s complaint/illness did not fit within the scope of the guidelines, nurse providers contacted an on-call physician for guidance and correspondingly adjusted the assessment/plan. Given that the study area had a low rate of pneumococcal vaccination^24^ and high rates of pneumococcal infection in children with acute ARI^25^, amoxicillin was indicated for most children with ARI. The study occurred after the 2010 cholera outbreak^26^ and during the onset of the COVID-19 pandemic.^27^

For those participants that received a household delivery, the nurse conducted a paired in-person clinical exam identical in scope to the phone-based call center exam with the addition of objective vital sign and anthropomorphic measurements. The purpose of the in-person exam was to establish a reference standard to evaluate the accuracy of the clinical data obtained by phone at the call center. Temperature was measured using a Welch Allyn SureTemp Plus Electronic Thermometer (orally ≥1 year), weight using a Taylor 7042 digital scale and heart rate and oxygen saturation using an Edan H100B Handheld Oximeter. Nasal swabs were collected from patients with ARI or fever, and rectal swabs were collected from patients with acute diarrhea.

### Laboratory Procedures

Nasal swabs were placed in Copan’s Universal Transport Medium; rectal swabs were placed in RNAlater. Samples were stored at -20C. Sample collection was paused for SARS-CoV-2 precautions on March 19, 2020, and recommenced on August 28, 2020. Respiratory samples from this date until the study concluded were analyzed using BioFire COVID-19 Test v1.0 or Respiratory Panel 2.1. The Respiratory Panel tests for 18 viral agents and four bacterial agents; it does not test for *Streptococcus* spp. Rectal swabs were stored for future molecular studies of diarrheal disease targets^28 29^.

### Operational procedures

The operational workflow was as follows (Figure S2): (i) A parent whose child was experiencing an illness contacted the call center. (ii) A call center healthcare provider used paper clinical decision-support tools to triage the child as mild, moderate, or severe (life-threatening). If the triage was severe, the child was referred to the hospital. If transport to emergency care could not be accessed, the TMDS offered transport. If the triage was not severe, the provider gathered basic exam findings and medical history from the parent over the phone to generate an assessment and plan. (iii) If the child lived within the delivery zone, a TMDS provider and driver were dispatched to conduct an in-person clinical exam and transport medications/fluids to the child’s home, otherwise the family received consult alone. (iv) All families received a follow-up call.

#### Training

The provider type was a licensed Haitian nurse with medical oversight from a licensed Haitian physician. Nurses and delivery drivers received didactic and experiential training. Prior to launch, 8 nurses participated in 4 days of interactive lectures on the study protocols, clinical decision support tools, professionalism, human subjects research and dispatch technology. They continued training on standardized and improvised case simulations through the first month of operations. Nine motorcycle drivers received one half day training on operational procedures, safety, professionalism, human subjects research and dispatch technology. A clinical advisory committee reviewed cases throughout the study to monitor for guideline adherence and unanticipated clinical events. Opportunities for quality improvement were identified and conveyed in an adaptive manner to the nurses and physicians and, when appropriate, the IRB protocols were amended. Logistics were reviewed and adjustments were conveyed to the team.

#### Technology

Call intake was managed by Twilio Flex (Twilio Inc.) and dispatch was managed by Beacon (Trek Medics International Inc.). Calls placed to a local phone number were forwarded to Twilio Flex and on to the call center web-based user interface where they were placed into a queue. Incidents (household visits/deliveries) were created in Beacon to alert, track, and coordinate drivers.

#### Call center

During hours of operation (6pm-5am), the call center was staffed by two nurses, two on-call motorcycle taxi drivers and two on-call physicians. Incoming calls were screened by a call center nurse for enrollment eligibility. Once eligibility was established, the nurse used the clinical tools to triage cases as life threatening (severe) or not life-threatening (mild or moderate) and generate an assessment and treatment plan. A map with community demarcations and landmarks was used to determine if households were inside or outside of the delivery zone.

#### User fees

Medication/fluid deliveries were offered for a fee on a sliding-scale starting at 500 Gourdes (5 USD) down to zero. The fee was set at a level commensurate with daytime delivery and medication costs. The amount to be paid was established at the call center without coercion or impact on enrollment eligibility or care. Fees were used to assess families’ willingness to pay and minimize incentive to delay seeking daytime care from local providers to obtain free nighttime care from the TMDS.

#### Dispatch

The two call center nurses alternated conducting household visits. One or two drivers were dispatched to transport the nurse from the call center to the child’s home. The decision to dispatch one or two drivers depended on the time of night (after 9pm), distance, and the security situation. GPS coordinates of households and tracks of delivery routes were recorded with a Garmin GPSMAP 64. Driver response times, including intervals for arrival at household and incident duration were collected using Beacon.

#### Household visit

Upon arrival at the household, the nurse obtained written informed consent from parents of participants and written informed assent from participants 7 to 10 years. The nurse conducted a paired in-person clinical exam. The data points were used to triage and assess the case independent of the plan developed at the call center. Most cases not requiring medications or fluids were given oral rehydration solution (ORS) as a teaching opportunity for future illness.

#### Follow-up

Severe cases, and cases with unsuccessful deliveries, were contacted (maximum of 2 attempts) by phone within 24-hours and all participants were contacted (maximum of 3 attempts) at 10-days to ascertain disposition, if additional care was sought, and qualitative feedback about the service.

### Clinical outcomes

Data were collected by Haitian providers on paper CRFs and digitized into REDCap. Ages were recorded in units of months from 0 to <2 years and units of years from 2 to 10 years. Clinical metrics were analyzed to describe the distribution of severity, complaints, disease type, medication/fluid prescriptions, and laboratory findings. Disease types were defined as follows: (i) Fever without source, (ii) ARI defined as cough with fever (excluded diarrhea), (iii) Diarrhea without ARI, (iv) ARI and diarrhea, and (v) ‘other disease type’. Sources of infection were defined as ARI, diarrhea, pain with urination, bacterial skin infection, scabies, vaginal discharge, and ear infection.

Clinical safety of the TMDS was assessed by conducting paired exams at the call center and at the household, and 10-day follow-up calls with all participants. Outcome measures were percentages of a given severity stratum (mild, moderate, severe) at the call center and at the household, as well as clinical status at the 10-day follow-up. Guideline deviations were analyzed by chart reviews performed by two independent Haitian physicians; conflicts between physician reviews were resolved on a case-by-case basis with discussion between the two physicians. Chart reviews focused on case severity, prescription of an antibiotic when not indicated, failure to prescribe an indicated antibiotic, and/or failure to identify a danger sign.

### Feasibility metrics

Feasibility metrics were analyzed and categorized according to the following conventions: demand, implementation, integration and acceptability.^30^ These outcomes were assessed using enrollment, time to delivery, additional care sought, and participant feedback data.

### Statistical approach

Repeat participants were considered unique cases if subsequent calls occurred 30 days or greater after the initial call. Case severity of bacterial skin infections were miscategorized during 14 months of the 16-month long study because of an error in training. The error was corrected *post hoc* (mild to moderate) prior to data analyses. Cases were categorized into disease types. Clustering attributed to repeat patient participants (>30 days between calls), repeat adult parent/guardian callers, and call center nurses was not considered. Detailed analyses on congruence between call center and in-person exams as a safety outcome will be presented in a concurrent publication.

Data were described by proportions for categorical variables and medians with quartiles for continuous variables. The Cochran-Mantel-Haenszel test was used to compare case severity by age, median age by disease type, follow-up care sought by case severity and feedback on the service by household visit status. Statistical significance was defined at α <0.05%. There is missingness due to the absence of household visit data for severe cases and loss to follow-up at 10-days. Analyses were completed in R version 4.1.1 and R studio version 2021.9.0.351 (R Foundation for Statistical Computing)^31^ and Statistical Analysis Software (SAS Institute) version 9.4.

## RESULTS

### Characteristics

A total of 391 cases were enrolled from September 9^th^, 2019 to January 19^th^, 2021; 46% (181) were female. The median age was 23 months; 7% (27) were less than 2 months and 43% (169) were between two months and less than two years (Table 1).

**Table 1.** Characteristics of enrolled cases

|  | Total | With household visit |
| --- | --- | --- |
| All cases | 391<sup>a</sup> | 347 |
| Age; median mo (Q1-Q3)<sup>b</sup> | 23 (9-48) | 24 (10-48) |
| <2 mo | 27 (7%) | 20 (6%) |
| 2 mo to <2 yrs | 169 (43%) | 150 (43%) |
| 2 yrs to <5 yrs | 121 (31%) | 112 (32%) |
| 5 yrs to 10 yrs | 74 (19%) | 65 (19%) |
| Sex; Female | 181 (46%) | 163 (47%) |

<sup>a</sup> Nine patients were repeat cases within 30 days, 68 were repeat cases within 323 days.
<sup>b</sup> (Q1-Q3), quarter 1 to quarter 3.

### Clinical Outcomes

The majority of cases were categorized as mild at both the call center (73%; 278) and household (79%; 268; Table 2). At the call center, 6% (24) of cases were identified as severe and referred to emergency care. Two percent (3 mild, 3 moderate) of cases triaged as not severe at the call center were re-categorized as severe at the household and referred to emergency care. At the call center, cases <2 months of age had the highest proportion of severe cases (23%; 6); case severity was associated with age (p=0.015). At the household cases <2 months had the highest proportion of severe cases (16%; 3); age was not associated with severity (p=0.082). The most common chief complaints at the call center were fever (43%; 59) and respiratory problems (18%; 67; Figure S3). By disease type at the household, ARI, diarrhea (without ARI), and fever represented 25% (84), 14% (47), and 11% (39) of cases respectively (Table 3). The median age of cases with diarrhea was less than other disease types at both the call center and household. ORS, paracetamol, and amoxicillin were provided to 81% (274), 59% (200) and 33% (112) of cases, respectively. Among cases with fever without a source, 15% (6) received an antibiotic and were all over 2 years (Table S1). Among cases with ARI (without diarrhea), 100% (84) received an antibiotic. Among cases with diarrhea (without ARI), 92% (43) received ORS and 19% (8) received an antibiotic; for cases between 2 months and less than 5 years, 53% (18) received zinc. Among cases with ARI and diarrhea, 100% (10) received an antibiotic, 100% (10) received ORS; for cases between 2 months and less than 5 years, none received zinc.

**Table 2.** Distribution of case severity

|  | At call center |  |  |  | At household |  |  |  |
| --- | --- | --- | --- | --- | --- | --- | --- | --- |
|  | Mild | Moderate | Severe | p<sup>b</sup> | Mild | Moderate | Severe | p<sup>b</sup> |
| All cases | 278 (73%) | 80 (21%) | 24 (6%) | --- | 268 (79%) | 64 (19%) | 6 (2%) | --- |
| Age; median mo (Q1-Q3)<sup>a</sup> | 24 (9-48) | 22 (12-48) | 12 (2-22) | 0.015 | 24 (10-48) | 23 (7-48) | 21 (10-33) | 0.082 |
| <2 mo | 16 (62%) | 4 (15%) | 6 (23%) | --- | 14 (74%) | 2 (10%) | 3 (16%) | --- |
| 2 mo to <2 yrs | 114 (70%) | 38 (23%) | 12 (7%) | --- | 109 (75%) | 34 (24%) | 2 (1%) | --- |
| 2yrs to <5 yrs | 94 (80%) | 20 (17%) | 4 (3%) | --- | 96 (87%) | 13 (12%) | 1 (1%) | --- |
| 5yrs to 10 yrs | 54 (73%) | 18 (24%) | 2 (3%) | --- | 49 (77%) | 15 (23%) | 0 (0%) | --- |

<sup>a</sup> (Q1-Q3), quarter 1 to quarter 3
<sup>b</sup>. p value of the Cochran-Mantel-Haenszel test (Nonzero Correlation) of association between severity and age.

**Table 3.**
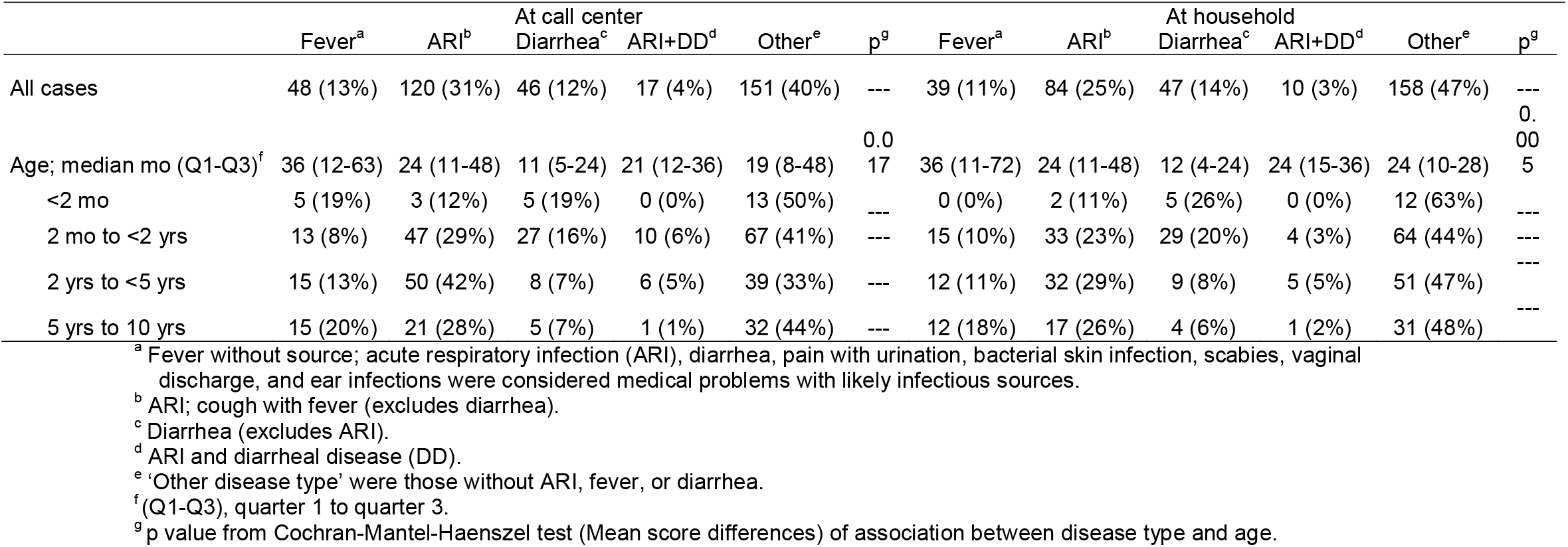
Distribution of disease type

Among patients with ARI or fever without source, whose samples were tested using the full respiratory panel, 88% (53) were positive for one or more pathogen and most pathogens identified were viruses (85%; 45). The three most common viruses detected were: Human Rhinovirus/Enterovirus (43%; 26), Respiratory Syncytial Virus (20%; 12), and Parainfluenza Virus 4 (10%; 6; Table S2). Five percent (4) were positive for SARS-CoV-2. Bacterial pathogens detected were *Bordetella parapertussis* (5%; 3), *Bordetella pertussis* (3%; 2), *Chlamydia pneumoniae* (3%; 2) and *Mycoplasma pneumoniae* (2%; 1).

At the 10-day follow-up call, 90% (348) of participants were reached. The distribution of clinical status was: 95% (329) were “improved” or “recovered”, 5% (18) were “the same”, and less than 1% (1) were “worse” (Table 4). Among those with severe conditions, 100% (24) of cases reported their condition as “improved” or “recovered”. Among the subset of cases who received a 24-hour follow-up call, 89% (39) reported their condition as “improved” or “recovered” at 24-hours. One severe adverse event occurred that involved a female patient less than 60 days old with a history of intermittent stooling, ‘weak cry’ and poor breastfeeding. The life-threatening status of the patient was not identified at the call center and the child died of unknown cause during transit to the household. The case was reported to the IRBs with an associated verbal autopsy.

**Table 4.** Clinical status at 10 days and additional care sought in the follow-up period

| Follow-up | Clinical status at follow-up |  |  | Care sought during the follow-up period |  |  |  | p<sup>b</sup> |
| --- | --- | --- | --- | --- | --- | --- | --- | --- |
|  | Improved/Recovered | Same | Worse | Hospital | Clinic | Other | None |  |
| All at ten days<sup>a</sup> | 329 (95%) | 18 (5%) | 1 (<1%) | 21 (6%) | 55 (16%) | 6 (2%) | 265 (76%) | <0.001 |
| Mild | 238 (94%) | 14 (6%) | 0 (0%) | 9 (3%) | 33 (13%) | 4 (2%) | 205 (82%) | --- |
| Moderate | 67 (93%) | 4 (6%) | 1 (1%) | 5 (7%) | 9 (12%) | 2 (3%) | 56 (78%) | --- |
| Severe | 24 (100%) | 0 (0%) | 0 (0%) | 7 (29%) | 13 (54%) | 0 (0%) | 4 (17%) | --- |
| Subset at 24-hours | 39 (89%) | 4 (9%) | 1 (2%) | 5 (11%) | 14 (32%) | 5 (11%) | 20 (46%) | --- |
| Mild | 10 (84%) | 1 (8%) | 1 (8%) | 0 (0%) | 2 (17%) | 2 (17%) | 8 (66%) | --- |
| Moderate | 8 (89%) | 1 (11%) | 0 (0%) | 1 (11%) | 3 (33%) | 0 (0%) | 5 (56%) | --- |
| Severe | 21 (91%) | 2 (9%) | 0 (0%) | 4 (17%) | 9 (39%) | 3 (13%) | 7 (31%) | --- |

<sup>a</sup> Includes cases in the 24-hour subset.
<sup>b</sup> p value from Cochran-Mantel-Haenszel test (Mean score differences) to compare care seeking during 10 day follow-up period by case severity.

Independent chart reviews found that 86% (327) of cases at the call center and 82% (275) of cases at the household were adherent to the guidelines (Table S3). Regarding deviations, mis-categorization between mild and moderate severity was the most common type of deviation at both the call center and household, occurring in 13% (48) and 10% (35) of cases respectively; 73% (35) and 89% (31) of this deviation type concerned bacterial skin infections. A missed danger sign occurred in <1% (1) of cases at the call center and 3% (10) of cases at the household. Prescription of antibiotics when not indicated occurred in 1% (5) and 5% (16) of cases and failure to prescribe an antibiotic occurred in 1% (2) and <1% (1) of cases at the call center and household respectively.

### Feasibility Metrics

#### Demand

The call center received 3,216 incoming calls from 1,122 unique phone numbers, of which 505 were received during operational hours (1.0 per night). Of these, 77% (391) of callers met inclusion criteria and were enrolled (Figure 1; 0.8 per night). Of the 391 enrolled cases, 89% (347) received a household visit. Twenty seven percent (106) of cases were repeat patients; 2% (9) were repeat patients within 30 days and thus excluded from sub-analyses. Twenty-nine percent (117) of cases involved repeat callers.

**Figure 1.**
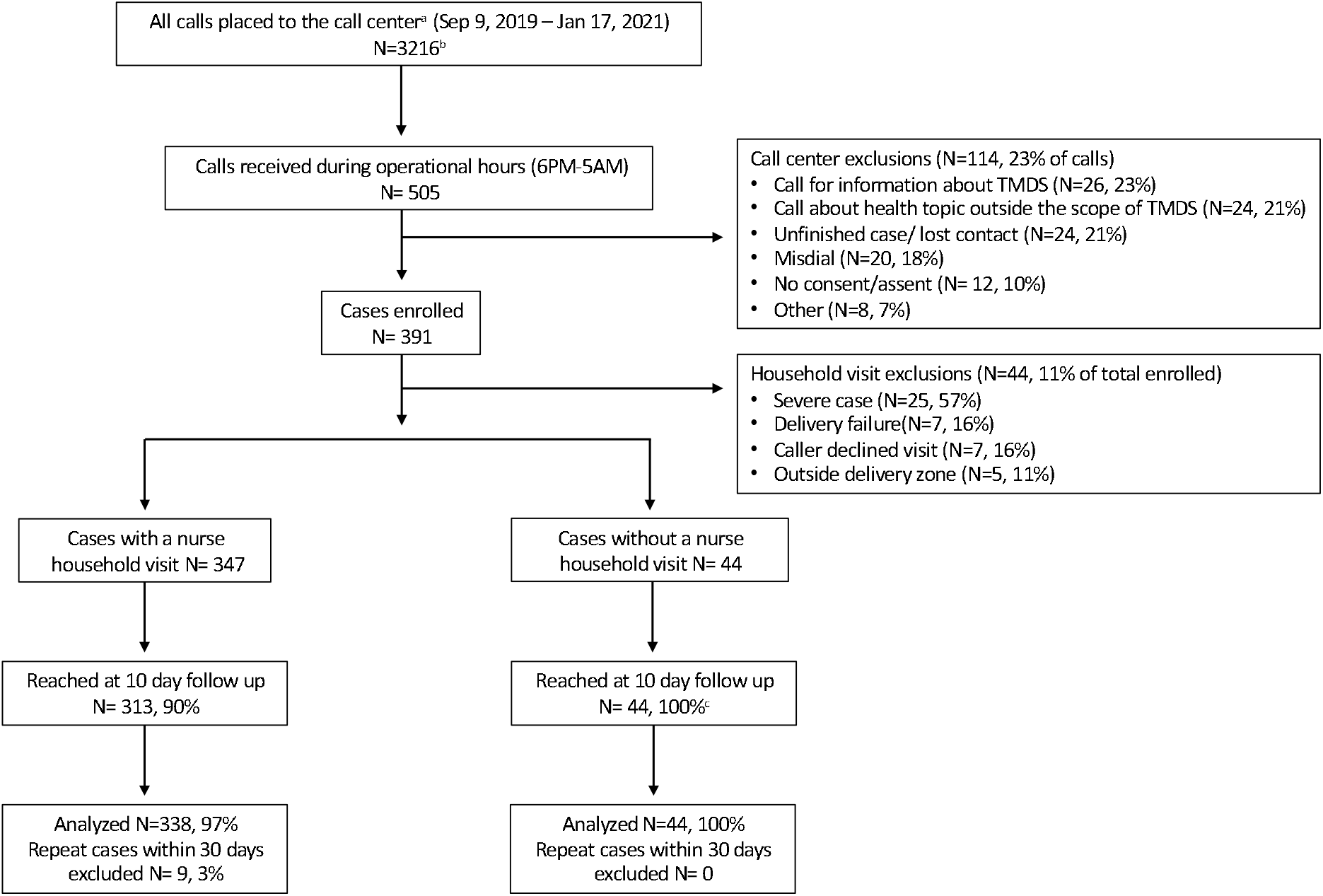
Diagram of case enrollment, reasons for exclusion, loss to follow up and inclusion in data analysis. ‘a’ = filtered to remove calls from employee phone numbers. ‘b’ = 1122 unique phone numbers. ‘c’ = cases not reached at 10 days were excluded because of no consent.

#### Implementation

The median phone consultation length was 20 minutes. The median time to arrival at the participant’s home was 73 minutes, with significant missingness due in part to connectivity problems. The median workflow duration per case, from the time the call was placed until the nurse returned to the call center from the household, was 114 minutes. Eleven percent (44) of cases did not receive a delivery and logistical challenges were the cause for 16% (7) of these instances.

#### Integration

At the 10-day follow-up, cases triaged as mild or moderate were less likely to have sought follow-up care than cases triaged as severe (p<0.001; Table 4). While the sample size was small, more cases triaged as severe sought referred care at a clinic (65%, 13) than a hospital (35%, 7) despite recommendations for hospital level care.

#### Acceptability

Among cases that received a household visit, greater than 99% (302) of callers rated the overall service as “good” or “great” (Table S4). Greater than 99% (293) of callers rated the delivery portion of the service as “good” or “great”. Greater than 99% (302) of callers responded that they would utilize the TMDS again. Regarding willingness to pay, 26% (89) contributed the full amount, another 22% (73) contributed partial payment and 99% (291) stated the price point was appropriate. Feedback on overall impression of the call center was significantly different between cases that did and did not receive a household visit (p<0.001). Cases without a visit were less likely to rate the service as great (63%; 27) than those who received a visit; (90%; 273).

## DISCUSSION

This study sought to evaluate the clinical safety and feasibility of implementing a TMDS for children at night in the resource limited setting of Haiti. From a clinical perspective, most cases were triaged as mild or moderate. Among the small percentage that were triaged as severe, the majority sought referred care. The percentage of cases initially triaged as not severe, yet were found to be severe at the household, was low. The majority of families reported the child’s condition as “improved” or “recovered” at follow-up. From a feasibility perspective, the duration of phone consultation and household deliveries were collectively completed well under our goal of two hours. Nearly all families rated the TMDS as “good” or “great” and repeat callers were common. Taken together, these findings suggest the TMDS was a feasible healthcare delivery model with high rates of improved clinical status.

With respect to clinical safety, this study was not designed as a clinical trial to evaluate morbidity and mortality outcomes. However, clinical safety metrics were enumerated. These metrics included case distributions, comparisons of severity at the call-center against the in-person reference standard at the household, clinical status at follow-up, as well as independent review of guideline adherence/deviations. The clinical approach was set based on the distributions of patient age and diseases found in the INACT1 needs assessment as well as rates documented in national^21 32^ and international^33^ resources. The case distributions (ages, complaints, disease types), or later by laboratory diagnostics, matched well with the expectations, and corresponding clinical decision support tools, set *a priori*. During the study some minor adjustments were made to the tools, however, major adjustments were not needed.

The distribution of disease severity aligned with the objectives of the TMDS as a pre-emergency service. The distribution of severity suggests that the messaging via print and radio advertisement was interpreted as intended. Among mild and moderate cases, the lower rates of seeking additional care suggests that the clinical problem was sufficiently addressed which is supported by the high rates of satisfaction at the 10-day follow-up. The successful engagement with the existing healthcare infrastructure for severe cases was reassuring, despite some seeking care the next day. Lastly, one in fifty cases were triaged as not severe yet found to be severe at the household. While this rate is low, future iterations of the TMDS will prioritize reducing this rate further.

The TMDS pilot model did have drawbacks. First, infants less than 2 months were most often triaged as severe at both the call center and household. Given the extreme vulnerability of these patients additional efforts to facilitate emergency care access at night is needed. Second, the medication supply was limited in the study area. For example, nodular scabies infections with secondary bacterial infection were common and resulted in high rates of oral antibiotics and topical scabies treatment; these cases would likely have benefitted from oral ivermectin over topical treatment, but ivermectin was not available. Similarly, there was disruption in zinc access for the supportive management of diarrheal disease. Third, the study area was largely unvaccinated for pneumococcus which resulted in high rates of oral antibiotic use for ARI out of empirical concern for bacterial pneumonia. To reduce rates of oral antibiotic usage, public health campaigns on scabies mitigation and pneumococcus vaccination might have significant benefit. Lastly, the laboratory findings also revealed causative agents of ARI in which amoxicillin would not be effective (e.g. *Mycoplasma pneumoniae* and *Bordetella pertussis*).

There was one known occurrence of a severe adverse event (SAE; death) out of 348 respondents (0.3%); per IRB evaluation, the death did not have a causative association with the TMDS. In response to the SAE, measures were enacted that included re-training on how to query the ability/inability to drink and assess dehydration, recognize abnormally normal vital signs in an ill pediatric patient and follow-up with clarifying questions at the level of the call center. The SAE rate of 0.3% is lower than the reported national under 5 mortality rate of 0.63% (63/1000 live births)^20^ and may serve as a safety effect size for future studies. In addition to the congruence analyses reported on triage, more granular analyses of the paired call center and household exams will be presented in a subsequent publication to identify additional areas for quality improvement and decision support tool iteration.

With respect to feasibility, we focused primarily on operational metrics. The phone consultation duration of 20 minutes was reasonable in the context of staff capacity with paper decision support tools, limited cell minutes and the battery life of callers’ phones. Despite limited data, the median time to arrival at the household (73 minutes) was well below the target of two hours; this target, coined the ‘silver two hours’, was based on an empiric assumption that the call center assessment of a non-severe case would hold for two hours. The median duration to complete the workflow per case was 114 minutes which can serve as a standard for scaling the TMDS. The drivers were able to locate the majority of households at night despite a lack of an address system, well paved roads or streetlights; this speaks to the versatility and value of local expertise among the drivers. While rare (2%), delivery failures were due to rain, insecurity, and loss of cellular contact with the caller. Acceptability was demonstrated by a high proportion of callers rating the TMDS as ‘good’ or ‘great’ and willing to utilize the TMDS in the future. Nearly all callers considered the price point as ‘appropriate’ yet only one quarter made a payment in full. However, half of callers did contribute which demonstrates they valued the service. Willingness to pay and models for payment (set fee, fee with waiver, sliding scale) are important factors in developing innovative sustainable healthcare solutions that do not undercut existing health access points. Literature on what is the ‘best’ approach varies and is often conflicted.^34-36^

These findings should be viewed within the context of the study limitations: (i) The methods of advertising varied over the course of the study to avoid overwhelming the nascent TMDS and adjust to political unrest, economic instability, and the COVID-19 pandemic. These factors likely influenced participant characteristics. (ii) Clinically, the lack of a malnutrition screen during the phone consultation meant there was no opportunity for nurse providers to identify severe malnutrition as a danger sign at the call center. This may have resulted in a less effective triage at the call center; four severely malnourished children were identified at household visits. (iii) Connectivity problems resulted in failures to reach households, complete phone consultations and phone follow-ups. These challenges lengthened quantitative operational metrics. (iv) Finally, the delivery zone was limited to a 5 km radius surrounding the call center where TMDS staff had a high level of familiarity with the local community, geography, and landmarks. It is unclear how important staff knowledge was in the successful implementation of the TMDS. Therefore, it is unknown how the TMDS would fair if a central call center serviced a geographically distant population.

## CONCLUSIONS

This study demonstrated that the TMDS was a feasible healthcare delivery model with high rates of improved clinical status at 10-days. Future efforts to evaluate scalability of the TMDS will include a change to the workflow and guidelines to provide mild cases with consult and delivery alone, and moderate cases with consult, delivery and a nurse visit, as well as evaluation in diverse settings.

## Data Availability

All data produced in the present study are available upon reasonable request to the authors.

## Acknowledgements

We are grateful to the participation of the patients and their parents/guardians. We also thank the field team who collected the data for this study. We are grateful to Randy Autrey and Krista Berquist for their administrative support, as well as Glenn Morris at the Emerging Pathogens Institute and Desmond Schatz interim Chair of the Department of Pediatrics at the University of Florida for their ongoing support, as well as guidance from the Ministry of Public Health and Population (Ministère de la Santé Publique et de la Population - MSPP). Map data are copyrighted by OpenStreetMap contributors and available from https://www.openstreetmap.org.

## Co-Author Contact Information

Molly B. Klarman

Departments of Pediatrics and Environmental and Global Health, University of Florida, Gainesville, FL, USA

Katelyn E. Flaherty

Departments of Emergency Medicine and Environmental and Global Health, University of Florida, Gainesville, FL, USA

Xiaofei Chi

Department of Health Outcomes and Biomedical Information, University of Florida Gainesville, FL, USA

Youseline Cajsuma

Departments of Pediatrics and Environmental and Global Health, University of Florida, Gainesville, FL, USA

Lerby Exantus

Université d’État d’Haiti-Faculté de Médecine et de Pharmacie, Port-au-Prince, Haiti

Jason Friesen

Trek Medics International. Washington, DC, USA

Valery M Beau de Rochars

Department of Health Services Research, Management and Policy, College of Public Health and Health Professions, University of Florida, Gainesville, FL, US

Chantale Baril

Université d’État d’Haiti-Faculté de Médecine et de Pharmacie, Port-au-Prince, Haiti

Matthew J. Gurka

Department of Health Outcomes and Biomedical Informatics, University of Florida Gainesville, FL, USA

Torben K. Becker,

Departments of Emergency Medicine and Environmental and Global Health, University of Florida, Gainesville, FL, USA

Eric J. Nelson

Departments of Pediatrics and Environmental and Global Health, University of Florida,

Gainesville, FL, USA

## References

1. United Nations Development Programme. Sustainable Development Goals: @undp; 2016 [Available from: https://www.undp.org/content/undp/en/home/sustainable-development-goals.html accessed March 20 2020.

2. The World Health Organization, The World Bank. Tracking Universal Health Coverage: 2017 Global Monitoring Report. Washington, D.C., 2017.

3. Peters DH, Garg A, Bloom G, et al. Poverty and access to health care in developing countries. Ann N Y Acad Sci 2008;1136:161–71. doi: 10.1196/annals.1425.011 [published Online First: 20071022]

4. Roark RF, Shah BR, Udayakumar K, et al. The need for transformative innovation in hypertension management. Am Heart J 2011;162(3):405–11. doi: 10.1016/j.ahj.2011.06.010 [published Online First: 20110811]

5. Bright T, Felix L, Kuper H, et al. A systematic review of strategies to increase access to health services among children in low and middle income countries. BMC Health Serv Res 2017;17 doi: 10.1186/s12913-017-2180-9

6. World Health Organization and UNICEF. Ending Preventable Child Deaths from Pneumonia and Diarrhoea by 2025. The integrated Global Action Plan for Pneumonia and Diarrhoea (GAPPD), 2013.

7. Temesgen ZM, DeSimone DC, Mahmood M, et al. Health Care After the COVID-19 Pandemic and the Influence of Telemedicine. Mayo Clinic Proceedings 2020;95(9):S66–S68. doi: 10.1016/j.mayocp.2020.06.052

8. Smith AC, Thomas E, Snoswell CL, et al. Telehealth for global emergencies: Implications for coronavirus disease 2019 (COVID-19). Journal of Telemedicine and Telecare 2020;26(5):309–13. doi: 10.1177/1357633X20916567

9. Williams S, Hill K, Xie L, et al. Pediatric Telehealth Expansion in Response to COVID-19. Frontiers in pediatrics 2021;9:642089–89. doi: 10.3389/fped.2021.642089

10. Galagali PM, Ghosh S, Bhargav H. The Role of Telemedicine in Child and Adolescent Healthcare in India. Current Pediatrics Reports 2021;9(4):154–61. doi: 10.1007/s40124-021-00253-w

11. Karari C, Tittle R, Penner J, et al. Evaluating the uptake, acceptability, and effectiveness of Uliza! clinicians’ HIV hotline: a telephone consultation service in Kenya. Telemed J E Health 2011;17(6):420–6. doi: 10.1089/tmj.2010.0220 [published Online First: 2011/05/27]

12. Alam M, Banwell C, Olsen A, et al. Patients’ and Doctors’ Perceptions of a Mobile Phone-Based Consultation Service for Maternal, Neonatal, and Infant Health Care in Bangladesh: A Mixed-Methods Study. JMIR Mhealth Uhealth 2019;7(4):e11842. doi: 10.2196/11842 [published Online First: 2019/04/23]

13. NowRX Telehealth. Telehealth the way it should be [Available from: https://nowrx.com/telehealth/ accessed December 15 2021.

14. National Health Security Office. Postal medicine delivery plays key role in telemedicine [Available from: http://eng.nhso.go.th/view/1/home/Postal-medicine-delivery-plays-key-role-in-telemedicine/230/EN-US accessed December 14 2021.

15. Rockethealth Telemedicine Laboratory Pharmacy, 2021 [Available from: https://www.rockethealth.shop/ accessed December 14 2021.

16. Klarman M, Schon J, Cajusma Y, et al. Opportunities to catalyse improved healthcare access in pluralistic systems: a cross-sectional study in Haiti. BMJ Open 2021;11(11):e047367. doi: 10.1136/bmjopen-2020-047367

17. Gage AD, Leslie HH, Bitton A, et al. Assessing the quality of primary care in Haiti. Bull World Health Organ 2017;95(3):182–90. doi: 10.2471/blt.16.179846

18. Tansley G, Schuurman N, Amram O, et al. Spatial Access to Emergency Services in Low-and Middle-Income Countries: A GIS-Based Analysis. PLOS ONE 2015;10(11):e0141113. doi: 10.1371/journal.pone.0141113

19. World Health Organization. World Health Statistics 2015. Luxemborg, 2015.

20. United Nations Inter-agency Group for Child Mortality Estimation. Levels and Trends in Child Mortality, 2020.

21. Vinekar K, Schaad N, Ber Lucien MA, et al. Hospitalizations and Deaths Because of Respiratory and Diarrheal Diseases Among Haitian Children Under Five Years of Age, 2011-2013. Pediatr Infect Dis J 2015;34(10):e238–43. doi: 10.1097/INF.0000000000000805

22. MINISTÈRE DE LA SANTÉ PUBLIQUE ET DE LA POPULATION. RAPPORT STATISTIQUE RÉPUBLIQUE D’HAÏTI MINISTÈRE DE LA SANTÉ PUBLIQUE ET DE LA POPULATION 2016. Port au Prince, Haiti, 2017.

23. Global burden of 369 diseases and injuries in 204 countries and territories, 1990-2019: a systematic analysis for the Global Burden of Disease Study 2019. Lancet 2020;396(10258):1204–22. doi: 10.1016/s0140-6736(20)30925-9

24. WHO and UNICEF. Haiti: WHO and UNICEF estimates of immunization coverage: 2019 revision.

25. Kim YY, Lew JF, Keith B, et al. Acute Respiratory Illness in Rural Haiti - International Journal of Infectious Diseases. 2019 doi: doi:10.1016/j.ijid.2019.02.003

26. Lemoine JF, Boncy J, Filler S, et al. Haiti’s Commitment to Malaria Elimination: Progress in the Face of Challenges, 2010–2016. Am J Trop Med Hyg 2017;97(4 Suppl):43–8. doi: 10.4269/ajtmh.16-0902

27. Rouzier V, Liautaud B, Deschamps MM. Facing the Monster in Haiti. New England Journal of Medicine 2020;383(1):e4. doi: 10.1056/NEJMc2021362

28. Flaherty KE, Grembi JA, Ramachandran VV, et al. High-throughput low-cost nl-qPCR for enteropathogen detection: A proof-of-concept among hospitalized patients in Bangladesh. PLOS ONE 2021;16(10):e0257708. doi: 10.1371/journal.pone.0257708

29. Grembi JA, Mayer-Blackwell K, Luby SP, et al. High-Throughput Multiparallel Enteropathogen Detection via Nano-Liter qPCR. Frontiers in cellular and infection microbiology 2020;10:351–51. doi: 10.3389/fcimb.2020.00351

30. Bowen DJ, Kreuter M, Spring B, et al. How we design feasibility studies. American journal of preventive medicine 2009;36(5):452–57. doi: 10.1016/j.amepre.2009.02.002

31. R: A Language and Environment for Statistical Computing [program]. Vienna, Austria: R Foundation for Statistical Computing, 2020.

32. Fene F, Ríos-Blancas MJ, Lachaud J, et al. Life expectancy, death, and disability in Haiti, 1990-2017: a systematic analysis from the Global Burden of Disease Study 2017. Revista panamericana de salud publica = Pan American journal of public health 2020;44:e136–e36. doi: 10.26633/RPSP.2020.136

33. Liu L, Oza S, Hogan D, et al. Global, regional, and national causes of under-5 mortality in 2000-15: an updated systematic analysis with implications for the Sustainable Development Goals. Lancet 2016;388(10063):3027–35. doi: 10.1016/S0140-6736(16)31593-8 [published Online First:2016/11/15]

34. Ponsar F, Tayler-Smith K, Philips M, et al. No Cash, No Care: How User Fees Endanger Health--Lessons Learnt Regarding Financial Barriers to Healthcare Services in Burundi, Sierra Leone, Democratic Republic of Congo, Chad, Haiti and Mali. International health 2011;3(2) doi: 10.1016/j.inhe.2011.01.002

35. Adams P. Health-care dynamics in Haiti. The Lancet 2010;376(9744):859–60. doi: https://doi.org/10.1016/S0140-6736(10)61396-7

36. Haar RJ, Naderi S, Acerra JR, et al. The livelihoods of Haitian health-care providers after the january 2010 earthquake: a pilot study of the economic and quality-of-life impact of emergency relief. International journal of emergency medicine 2012;5:13–13. doi: 10.1186/1865-1380-5-13

