## Supplementary_Materials for "Implementation of a pediatric telemedicine and medication delivery service in a resource-limited setting: A pilot study for clinical safety and feasibility"

**Text 3.** Formulary of available oral and topical medications and fluids………….....…………....21

**Figure S1**. Map of the call center, delivery zone, and delivery tracks

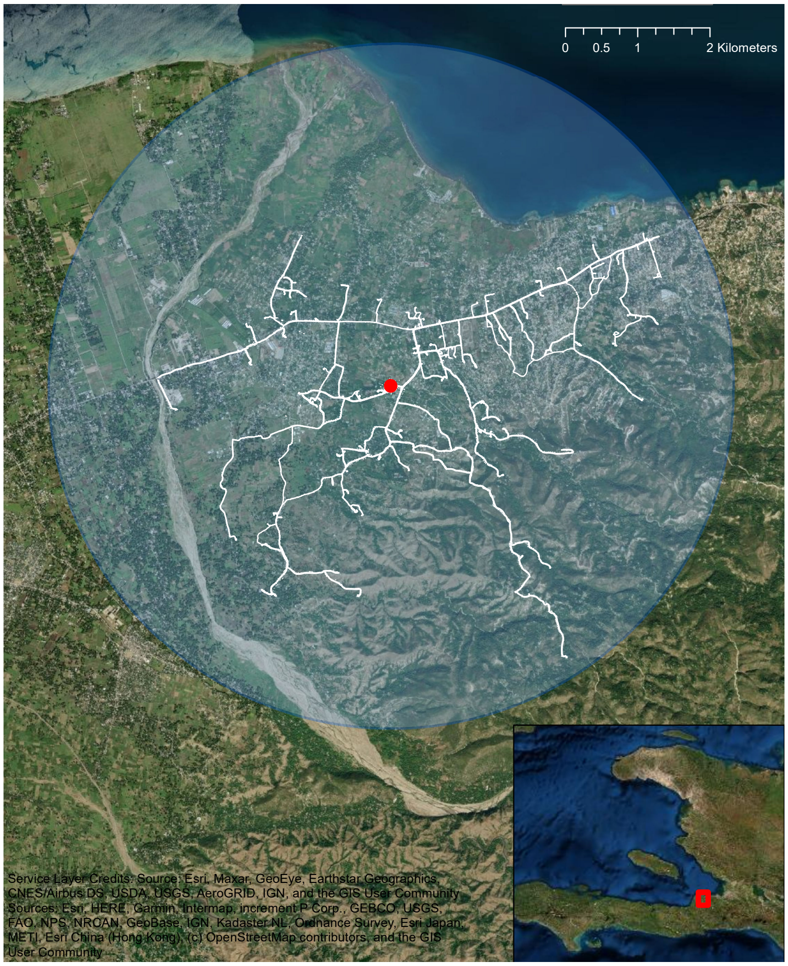

**Figure S1.** Map of the call center, delivery zone, and delivery tracks. Red circle = call center.

**Figure S2.** Telemedicine and delivery service workflow.

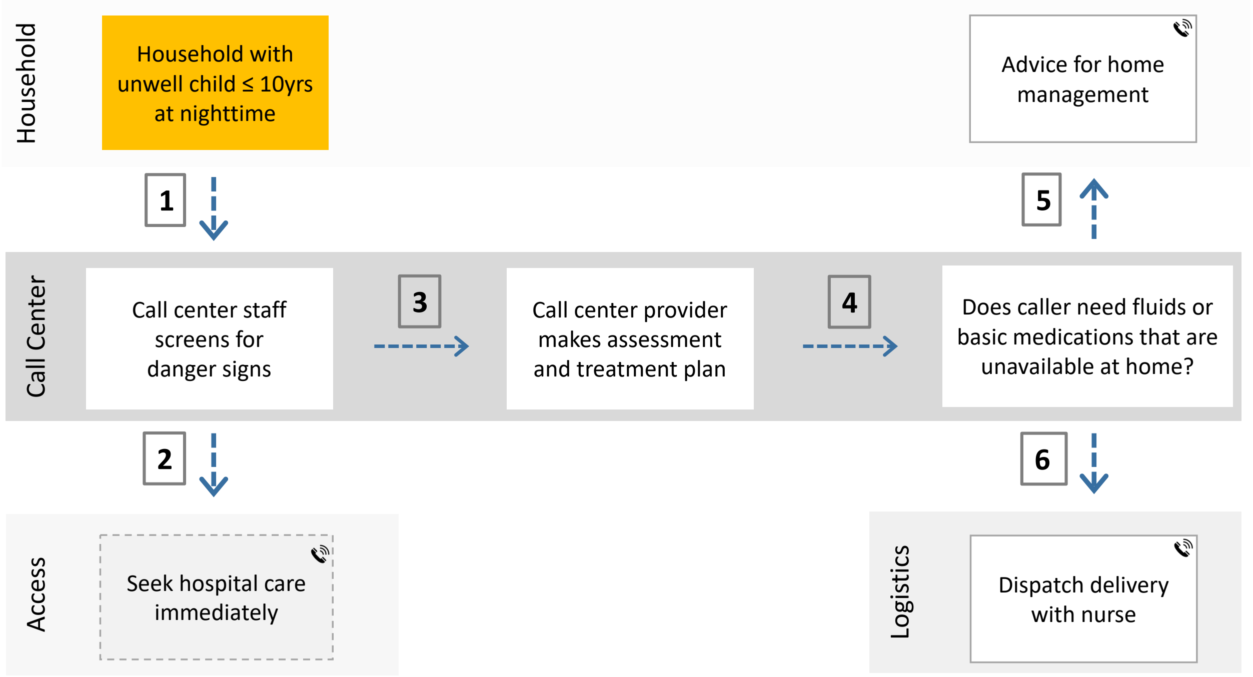

**Figure S2.** Telemedicine and delivery service workflow. (1) A parent contacts the call center. (2) A call center provider screens for danger signs; if present the child is referred to the hospital. (3) The provider gathers basic exam findings and medical history from the parent. (4) An assessment and treatment plan are generated. (5) If the child does not need medications/fluids or lives outside the delivery zone the family receives consult alone. (6) If the child lives within the delivery zone and needs medications/fluids, a TMDS provider and driver are dispatched to conduct clinical exam and transport items. All families received a follow-up call at 10 days. The original workflow included an automated message instructing callers experiencing an emergency to seek emergency care. The message was removed after 3 months to prevent callers from hanging up when not initially greeted by a live person. A version of this figure has been published previously[1] and is published here with permission from AJTMH.

1. Flaherty KE, Klarman MB, Cajusma Y, et al. A Nighttime Telemedicine and Medication Delivery Service to Avert Pediatric Emergencies in Haiti: An Exploratory Cost-Effectiveness Analysis. *The American Journal of Tropical Medicine and Hygiene.* 06 Apr. 2022 2022;106(4):1063-1071.

**Figure S3.** Distribution of medical complaints at the call center.

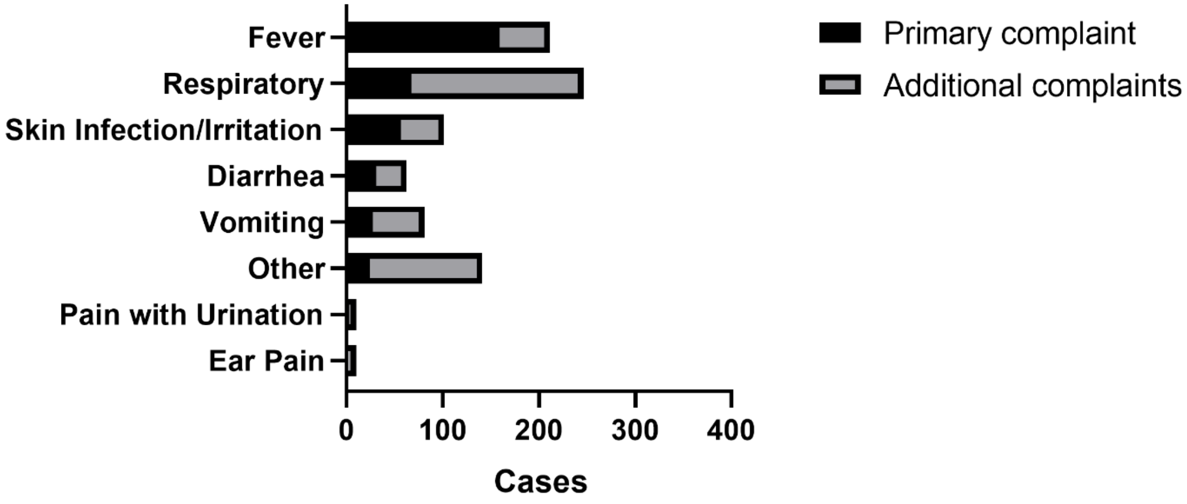

**Figure S3.** Distribution of medical complaints at the call center. Common ‘other complaints’ included headache, abdominal pain, and loss of appetite.

**Table S1.** Distribution of medications and fluids provided at household

| | Oral antibiotic | ORS | Zinc 2 months to <5 yrs |
|---|---|---|---|
| All cases with visit; N=338 | 162 (48%) | 274 (81%) | --- |
| Fever without source [a]; N=39 | 6 (15%) | 36 (92%) | --- |
| ARI[b]; N=84 | 84 (100%) | 81 (96%) | --- |
| Diarrhea[c]; N=47 | 9[d] (19%) | 43[e] (92%) | 18 (53%)[f] |
| ARI+Diarrhea; N=10 | 10 (100%) | 10 (100%) | 0 (0%)[g] |

[a] Fever without source; acute respiratory infection (ARI), diarrhea, pain with urination, bacterial skin infection, scabies, vaginal discharge, and ear infections were considered medical problems with likely infectious sources.

[b] ARI; cough with fever (excludes diarrhea).

[c] Diarrhea (excludes ARI).

[d] 1 case had bloody diarrhea, 1 had watery diarrhea and 7 were given an antibiotic for a different problem.

[e] 4 cases not receiving ORS were breastfeeding and <4 months; did not require ORS.

[f] 34 cases were within age range for zinc prescription

[g] 9 cases were within age range for zinc prescription

**Table S2**. Pathogens detected in respiratory samples

| Pathogens detected | Cases (N=60) |
|---|---|
| Human Rhinovirus/Enterovirus[a] | 26 (43%) |
| Respiratory Syncytial Virus | 12 (20%) |
| Parainfluenza Virus 4 | 6 (10%) |
| Adenovirus | 5 (8%) |
| Influenza B | 5 (8%) |
| Severe Acute Respiratory Syndrome Coronavirus 2 | 4 (5%)[b] |
| Parainfluenza Virus 2 | 4 (7%) |
| *Bordetella parapertussis* | 3 (5%) |
| Coronavirus NL63 | 2 (3%) |
| Coronavirus 229E | 2 (3%) |
| Human Metapneumovirus | 2 (3%) |
| Parainfluenza Virus 1 | 2 (3%) |
| Parainfluenza Virus 3 | 2 (3%) |
| *Bordetella pertussis* | 2 (3%) |
| *Chlamydia pneumoniae* | 2 (3%) |
| Coronavirus OC43 | 1 (2%) |
| Influenza AH1 | 1 (2%) |
| Influenza AH12009 | 1 (2%) |
| *Mycoplasma pneumoniae* | 1 (2%) |
| Coronavirus HKU1 | 0 |
| Influenza A | 0 |
| Influenza AH3 | 0 |

[a] One severe case.
[b] An additional 14 samples were analyzed for Severe Acute Respiratory Syndrome Coronavirus alone.

**Table S3.** Distribution of guideline adherence and deviations

| | At call center N=382 | At household N=337 |
|---|---|---|
| Cases fully adherent | 327 (86%) | 275 (82%) |
| Triage deviations | | |
| Miscategorized moderate as mild | 38[a] (10%) | 34[b] (10%) |
| Miscategorized mild as moderate | 10 (3%) | 1 (<1%) |
| Missed danger sign | 1 (<1%) | 10 (3%) |
| Antibiotic prescription deviations | | |
| Prescribed antibiotic when not indicated | 5 (1%) | 16 (5%) |
| Missed antibiotic prescription when indicated | 2 (1%) | 1 (<1%) |

[a] 35 deviations were due to systematic mis-categorization of secondary bacterial skin infections.
[b] 31 deviations were due to systematic mis-categorization of secondary bacterial skin infections.

**Table S4.** Participant feedback at 10 days

| | With household visit N=304 | Without household visit N=44 | p[a] |
|---|---|---|---|
| Overall impression of call center | | | <0.001 |
| Great | 273 (90%) | 27 (63%)[a] | --- |
| Good | 29 (10%) | 15 (35%) | --- |
| Okay | 1 (<1%) | 1 (2%) | --- |
| Overall impression of delivery | | | |
| Great | 272 (93%) | --- | --- |
| Good | 21 (7%) | --- | --- |
| Okay | 0 (<1%) | --- | --- |
| Would call again; Yes | 302 (> 99%) | 43 (100%) | --- |
| Fee payment | | | |
| Full | 89 (26%) | --- | --- |
| Partial | 73 (22%) | --- | --- |
| None | 173 (52%) | --- | --- |
| Fee amount | | | |
| Appropriate | 291 (99%) | --- | --- |
| Expensive | 3 (1%) | --- | --- |

[a] p value from Cochran-Mantel-Haenszel test (Mean score differences) to compare overall impression of the call center between cases with and without a household visit.

Text 1. Case Report Form

**INACT2 CASE REPORT FORM/ DECISION SUPPORT TOOL** **Patient ID __ -__ __ __ __ -__** ['P/X'-####A]

### IDENTIFICATION AND LOGISTICS

**IDENTIFICATION** **NURSE ID ___Initials _______**

| | | | |
|---|---|---|---|
| 1. __ __/ __ __/__ __<br>2C. Duration call<br>___ ___ min | 2A.Time: _ _ : _ _ am ☐ pm☐<br>2B.Repeat caller: ☐ No ☐ Yes<br>dd _ _/ mm _ _ | 3. < 2 years: __ __ months<br>≥ 2 years: __ __ years | 4. Location |
| 5. Caller Name (First/ Last) | | | 6. Primary Mobile<br>509 __ __ __ __ -__ __ __ __ |
| 7. Patient Name (First/ Last) | | 8. Sex: ☐ F ☐ M | 9. Secondary Mobile<br>509 __ __ __ __ -__ __ __ __ |
| 10A. Problem(s)<br>10B. Duration:<br>__ __ days | ☐ Fever #1 ☐ Cough/ Breathing #2 ☐ Vomit #3A<br>☐ Diarrhea #3B ☐ Ear Pain #4 ☐ Skin #5<br>☐ Pain with urine #6 ☐ Other #7: (free text) | | 11. Landmarks |

### 24-hour Follow Up

**☐ Need ☐ Not Needed**

**Required for RED patients and those referred to a clinic** **NURSE ID ___Initials _______**

| | | | |
|---|---|---|---|
| First call attempt | Time: _ _ : _ _ am ☐ pm☐ | dd _ _mm _ _yy _ _ | ☐ not completed ☐ completed |
| Second call attempt | Time: _ _ : _ _ am ☐ pm☐ | dd _ _mm _ _yy _ _ | ☐ not completed ☐ completed |
| 12. Condition | Current status: ☐ Well ☐ Sick/Improving ☐ Sick/Same ☐ Sick/Worsening ☐ Died ☐ Unknown | | |
| 13A. Care sought | Level of care patient first sought | | ☐ Hospital ☐ Clinic ☐ None ☐ Other:_ |
| 13B. Problem | If yes, what did they consider the main problem? | | ☐ Same ____(#) ☐ Different _______ |
| 13C. Intervention | If care sought, what was done? | | |
| 14. Patient location | Is the patient still receiving care? Where? | | ☐No (Home) Yes:☐Hospital ☐Other:_ |
| 15. Comments | | | |

### 10-day Follow Up

**Required for all patients** **NURSE ID ___Initials _______**

| | | | |
|---|---|---|---|
| First call attempt | Time: _ _ : _ _ am ☐ pm☐ | dd _ _mm _ _yy _ _ | ☐ not completed ☐ completed |
| Second call attempt | Time: _ _ : _ _ am ☐ pm☐ | dd _ _mm _ _yy _ _ | ☐ not completed ☐ completed |
| Third call attempt | Time: _ _ : _ _ am ☐ pm☐ | dd _ _mm _ _yy _ _ | ☐ not completed ☐ completed |
| 16. Condition | Current status: ☐ Well ☐ Sick/Improving ☐ Sick/Same ☐ Sick/Worsening ☐ Died ☐ Unknown | | |
| 17A. Care sought (1) | Level of care patient first sought | | ☐ Same as in 24 hour follow-up<br>☐ Hospital ☐ Clinic ☐ None ☐ Other:_ |
| 17B. Problem | If yes, what did they consider the main problem? | | ☐ Same ____(#) ☐ Different _______ |
| 17C. Intervention | If care sought, what was done? | | |
| 18A. Care Sought (2) | Was care sought a second time? | | ☐ Hospital ☐ Clinic ☐ None ☐ Other:_ |
| 18B. Problem | If yes, what did they consider the main problem? | | ☐ Same ____(#) ☐ Different _______ |
| 18C. Intervention | If care sought, what was done? | | |
| 19. Patient location | Is the patient still receiving care? Where? | | ☐ No (Home) Yes:☐Hospital ☐Other:_ |
| 20. Medication | Was medication given taken? ☐ Yes ☐ None prescribed ☐ No, why? | | |
| 21. Comments | | | |

**Feedback required for all patients**

| | | |
|---|---|---|
| 22. Price of service appropriate (only those that receive delivery). ☐ NA ☐ Yes ☐ Too expensive | | |
| 23. Overall impression of delivery service | | ☐ NA ☐ Great ☐ Good ☐ Okay ☐ Poor ☐ Bad |
| 24. Overall impression of call service | | ☐ Great ☐ Good ☐ Okay ☐ Poor ☐ Bad |
| 25. Would call again? | ☐ Yes ☐ No<br>If No, why? | 26. Additional feedback: |

#### CALL CENTER TRIAGE

NURSE ID___ Initials _______

**DANGER SIGNS** (danger sign **stop** and send to the hospital. Plan A. RED)

| | | |
|---|---|---|
| 27. General | Unresponsive (with stimulation) / lethargic | ☐ **YES** ☐ No |
| 28. Hydration | Able to drink / breastfeed<br>(effectively drank/breastfed without major vomiting) | ☐ **NO** ☐ Yes |
| 29. Neurologic | Has/does the child have a seizure (last 24 hours) | ☐ **YES** ☐ No |

**VITAL SIGNS (see vital sign table for reference)**

| | | |
|---|---|---|
| 30. Temp / fever | Under 1 year measured rectally (fever ≥38C / 100.4F)<br><br>Above 1 year orally (Fever ≥ 37.5C / 99.5F; uncorrected) | Fever: ☐YES (Subj), ☐ No (subj)<br>☐ Yes (Obj), ☐ No (obj)<br>Measurement: _ _ _ . _ ☐ C ☐ F<br>☐ Oral ☐ Axillary ☐ Rectal |
| 31A.Respiration Rate | Count chest rises in 15 seconds<br>• Under 1 year **>= 60** breaths/min, hospital<br>• 1 year or older **>=50**, hospital<br>• If < 2 yr and **<24** breaths/min, hospital<br>• If >= 2 yr and **<18** breaths/min, hospital | _ _ per 15 second x 4 for rpm<br>_ _ breaths/min<br>☐ Not obtained/ unreliable |
| 31B. Fast breathing | Breaths per minute:<br>Under 1 year >= 50,<br>1 year or older >= 40<br>Fast breathing could be a sign of pneumonia | ☐ Yes ☐ No<br>☐ Not obtained/ unreliable |
| 32. Heart Rate | Only for children less than 5 years, parent counts for 15 seconds with hand over left chest<br>• If **>= 180** beats/min, hospital (all ages)<br>• If < 2 yr and **<100** beats/minute, hospital<br>• If >= 2 yr and **<60** beats/minute, hospital | _ _ _ per 15 second X 4 for bpm<br>_ _ _ beats/min<br>☐ Not obtained/unreliable |

#### CALL CENTER PROBLEM SPECIFIC QUESTIONS

**Breathing problem / Cough #2.** **CONFIDENT**

| | | | | |
|---|---|---|---|---|
| 33. Head | Head goes up and down when breathing more than normal ('head bobbing') | ☐ YES | ☐ No ☐ Unknown | ☐ Yes ☐ No |
| 34. Nose | Do nostrils go in and out when breathing more than normal ('nasal flaring') | ☐ YES | ☐ No ☐ Unknown | ☐ Yes ☐ No |
| 35. Nose | Nasal discharge. Mucus is a sign of infection (a source) | ☐ YES | ☐ No ☐ Unknown | NA |
| 36. Mouth | Is there a cough | ☐ YES | ☐ No ☐ Unknown | NA |
| 37. Neck/Chest: | Chest indrawing showing ribs when breathing (retractions) more than normal | ☐ YES | ☐ No ☐ Unknown | ☐ Yes ☐ No |
| 38. Neck: Stridor | When calm, abnormal sound when breaths in (Stridor) that is different than the normal sounds | ☐ **YES** | ☐ No ☐ Unknown | ☐ Yes ☐ No |
| 39. Chest: Wheeze | When calm, wheeze when child breaths out (may be Asthma) that is different than the normal or nasal mucus sounds | ☐ YES | ☐ No ☐ Unknown | ☐ Yes ☐ No |
| If confident in # 36, 37, and 40, treat at hospital as red if all three are yellow. If confident in #41, treat at hospital. | | | | |

### CONTINUED CALL CENTER QUESTIONS

#### DEHYDRATION (all patients)

| | | |
|---|---|---|
| 40A. Urine (all ages) | Was there urine in the last 8 hours? | ☐ Yes ☐ No ☐ Unknown |
| 40B. Tears (<5 years) | Tears present? (with or without crying) | ☐ Yes ☐ No(do42-49)☐ Unknown |
| 41A. Vomiting **(#3A)** | Episodes in last 24 hours: ☐ None ☐ 1-2 ☐ 3-5 ☐ 6-11 ☐ 12 or more ☐ Unknown number | |
| 41B. Vomiting | Is vomit dark green vomit, If yes **Red** | ☐ Yes ☐ No ☐ Unknown |

#### DEHYDRATION (diarrhea patients) #3B

| | | | | CONFIDENT |
|---|---|---|---|---|
| 42A. General: ≥ 5 y | ☐ Well/Alert | --------BLANK-------- | ☐ Lethargic/ Unconscious **Red** | ☐ Yes ☐ No |
| 42B. General: < 5 y | ☐ Well/Alert | ☐ Restless/irritable | ☐ Lethargic/Unconscious **Red** | ☐ Yes ☐ No |
| 43. Eyes | ☐ Not Sunken | --------BLANK-------- | ☐ Sunken | ☐ Yes ☐ No |
| 44. Thirst | ☐ Normal | ☐ Drinks eagerly/thirsty | ☐ Not able to drink (effectively) | ☐ Yes ☐ No |
| 45. Skin pinch | ☐ Normal (<2sec) | ☐ Slow (2-3 sec) | ☐ Very slow (>3= sec) | ☐ Yes ☐ No |
| 46.Assess by scoring highest 2 findings | ☐ Green (<5%; No)<br>Score only those items marked as "Yes" for confident in the answer. | ☐ Yellow(5-10%;Some) | ☐ **Red**(>10%;Severe, hospital) | |

| | | | | |
|---|---|---|---|---|
| 47. Loose stools | Loose stools in last 24 hours: ☐ 1-2 ☐ 3-5 ☐ 6-11 ☐ 12 or more | | | |
| 48. Rice-water stool | Is it like rice-water (see plan for antibiotics) | ☐ Yes | ☐ No | ☐ Unknown |
| 49. Bloody stool | If yes, will require an antibiotic | ☐ Yes | ☐ No | ☐ Unknown |

#### EAR PAIN #4

| | | | | |
|---|---|---|---|---|
| 50. Puss from ear | If yes, antibiotics. If not, supportive care | ☐ Yes | ☐ No | ☐ Unknown |
| 51. Pain behind ear | Is the bone behind the ear painful when pressed, red, and swollen. Consider mastoiditis (red), typically follows an ear infection. | ☐ **Yes** | ☐ No | ☐ Unknown |

#### SKIN PROBLEM #5

| | |
|---|---|
| 52A. Type | ☐ Allergic (itchy with an 'exposure') ☐ Bacterial Infection (red, raised, warm, pain)<br>☐ Scabies (itchy small bumps) ☐ Other (e.g. viral rash): |
| 52B. If bacterial, size | ☐ Size of a coin ☐ Size of a hand ☐ Larger than a hand (hospital or morning follow-up) |

#### PAIN WITH URINATION #6

| | | | | |
|---|---|---|---|---|
| 53. Painful urination | If yes and there is a fever, suggests urine infection. | ☐ Yes | ☐ No | ☐ Unknown |

#### OTHER PROBLEMS #7 Ask questions that target the specific problem and gauge severity.

| |
|---|
| 54. Other problem |

#### PAST MEDICAL HISTORY

| | |
|---|---|
| 55. Past problems | |
| 56A. Medications:<br><br>56B. When was the last dose given? | 57. Allergies: ☐ No, ☐ Unknown, Yes: |

### HOUSEHOLD TRIAGE. NURSE ID___ Initials _______ ☐ REFUSED/ OUT OF AREA/ FAILED

**DANGER SIGNS** (danger sign **stop** and send to the hospital. Plan A. RED)

| | | |
|---|---|---|
| 58. General | Unresponsive (with stimulation) / lethargic | ☐ **YES** ☐ No |
| 59. Hydration | Able to drink / breastfeed<br>(effectively drank/breastfed without major vomiting) | ☐ **NO** ☐ Yes |
| 60. Neurologic | Has/does the child have a seizure (last 24 hours) | ☐ **YES** ☐ No |

**VITAL SIGNS (see vital sign table for reference)**

| | | |
|---|---|---|
| 61. Temp / fever | Under 1 year measured rectally (fever >= 38degC)<br>Above 1 year orally (Fever >= 37.5; uncorrected)<br>Do axillary temp on all (Fev.>= 37.5; uncorrected) | Fever: ☐ Yes (Obj), ☐ No (Obj)<br>☐ Oral ☐ Rect _ _ _ . _ ☐ C ☐ F<br>Axillary: _ _ _ . _ ☐ C ☐ F |
| 62A.Respiration Rate | Count chest rises in 15 seconds<br>• Under 1 year **>= 60** breaths/min, hospital<br>• 1 year or older **>=50** breaths/min, hospital<br>• If < 2 yr and **<24** breaths/min, hospital<br>• If >= 2 yr and **<18** breaths/min, hospital | _ _ per 15 second x 4 for rpm<br>_ _ breaths/min ☐ Not obtained |
| 62B. Fast breathing | Breaths/min: Under 1 year >= 50, 1 year or older >= 40. Fast breathing could be a pneumonia sign. | ☐ YES ☐ No ☐ Unsure |
| 63. Heart Rate<br>(pulse-ox or palpation) | • If **> 180** beats/min, hospital (all ages)<br>• If < 2 yr and **<100** beats/minute, hospital<br>• If >= 2 yr and **<60** beats/minute, hospital | _ _ _ per 15 second X 4 for bpm<br>_ _ _ beats/min ☐ Not obtained |

**VITAL SIGNS**

| | | |
|---|---|---|
| 64. Oxygen | If **< 90%** while awake, **stop** and direct to hospital | _ _ _ % |
| 65. Weight | Measure with minimal clothing (as appropriate) | _ _ _ . _ _ kg |
| 66. MUAC | Malnutrit: 2mo-6 mo if **<110mm**; 6mo-<5 yr**<115mm** | _ _ _ mm |

### HOUSEHOLD PROBLEM SPECIFIC QUESTIONS

**Breathing problem / Cough #2.**

| | | |
|---|---|---|
| 67. Head | Does head go up and down when breathing more than normal ('head bobbing') | ☐ YES ☐ No ☐ Unknown |
| 68. Nose | Do nostrils go in and out when breathing more than normal ('nasal flaring) | ☐ YES ☐ No ☐ Unknown |
| 69. Nose | Nasal discharge. Mucus is a sign of infection (a source) | ☐ YES ☐ No ☐ Unknown |
| 70. Mouth | Is there a cough | ☐ YES ☐ No ☐ Unknown |
| 71. Neck/Chest: | Chest indrawing showing ribs when breathing (retractions) more than normal | ☐ YES ☐ No ☐ Unknown |
| 72. Neck: Stridor | When calm, abnormal sound when breaths in (Stridor) that is different from normal | ☐ YES ☐ No ☐ Unknown |
| 73. Chest: Wheeze | When calm, wheeze when breaths out (may be asthma, noisy noise) that is different than the normal sounds. | ☐ YES ☐ No ☐ Unknown |
| 74. Chest: Stethoscope | When listening with a stethoscope, abnormal breaths sounds? | ☐ Normal ☐ Wheeze (out)<br>☐ Crackles ☐Other ☐Unknown |
| | | If more than two yellow signs, treat at hospital as red. |

### CONTINUED HOUSEHOLD QUESTIONS

#### DEHYDRATION (all patients)

| | | |
|---|---|---|
| 75A. Urine (all ages) | Was there urine in the last 8 hours? | ☐ Yes ☐ No ☐ Unknown |
| 75B. Tears (<5 years) | Tears present? (with or without crying) | ☐ Yes ☐ No (do 77-84) ☐ Unknown |
| 76A. Vomiting **(#3A)** | Episodes in last 24 hours: ☐ None ☐ 1-2 ☐ 3-5 ☐ 6-11 ☐ 12 or more ☐ Unknown number | |
| 76B. Vomiting | Is vomit dark green, If yes **Red** | ☐ Yes ☐ No ☐ Unknown |

#### DEHYDRATION (diarrhea patients) #3B

| | | | |
|---|---|---|---|
| 77A. General: > 5 y | ☐ Well/Alert | --------BLANK-------- | ☐ Lethargic/ Unconscious **Red** |
| 77B. General: < 5 y | ☐ Well/Alert | ☐ Restless/irritable | ☐ Lethargic/ Unconscious **Red** |
| 78. Eyes | ☐ Not Sunken | --------BLANK-------- | ☐ Sunken |
| 79. Thirst | ☐ Normal | ☐ Drinks eagerly/thirsty | ☐ Not able to drink (effectively) |
| 80. Skin pinch | ☐ Normal (<2sec) | ☐ Slow (2-3 sec) | ☐ Very slow (>3= sec) |
| 81.Assess by scoring highest 2 findings | ☐ Green (<5%; No) | ☐ Yellow (5-10%;Some) | ☐ **Red** (10%>; Severe, send to hospital) |

| | | |
|---|---|---|
| 82. Loose stool | How many stools in the last 24 hours | ☐ 1-2 ☐ 3-5 ☐ 6-11 ☐ 12 + |
| 83. Rice-water stool | Is it like rice-water (see plan for antibiotics) | ☐ Yes ☐ No ☐ Unknown |
| 84. Bloody stool | If yes, will require an antibiotic | ☐ Yes ☐ No ☐ Unknown |

#### EAR PAIN #4

| | | |
|---|---|---|
| 85. Puss from ear | If yes, antibiotics. If not, supportive care | ☐ Yes ☐ No ☐ Unknown |
| 86. Pain behind ear | Is the bone behind the ear painful when pressed, red, and swollen. This could be mastoiditis (red). | ☐ **Yes** ☐ No ☐ Unknown |

#### SKIN PROBLEM #5

| | |
|---|---|
| 87A. Type | ☐ Allergic (itchy with an 'exposure') ☐ Bacterial Infection (red, raised, warm, pain)<br>☐ Scabies (itchy small bumps) ☐ Other (e.g. viral rash): |
| 87B. If bacterial, size | ☐ Size of a coin ☐ Size of a hand ☐ Larger than a hand (hospital or morning follow-up) |

#### PAIN WITH URINATION #6

| | | |
|---|---|---|
| 88. Painful urination | If yes and there is a fever, suggests urine infection. | ☐ Yes ☐ No ☐ Unknown |

#### OTHER PROBLEMS #7 Ask questions that target the specific problem and gauge severity.

| |
|---|
| 89. Other problem |

#### PAST MEDICAL HISTORY

| | |
|---|---|
| 90. Past problems | |
| 91. Medications (current): | 92. Allergies: ☐ No, ☐ Unknown, Yes: |

### ASSESSMENT AND PLAN

#### CALL CENTER

| | | |
|---|---|---|
| 93. PLAN | ☐ **A. Mild** ☐ **B. Moderate** ☐ **C. Severe** (hospital) | |
| 94. Fever #1 (☐ not needed) | Paracetamol 15 mg/ kg every 6 hours as needed for fever. | |
| 95. Breathing/ Cough#2 (☐ not needed)<br><br>Note: It is ok to refill salbutamol/ albuterol for known asthmatics that are now without medication. | Pneumonia (fever and cough): Amoxicillin 40 mg/kg per dose by mouth twice daily for 5 days<br><br>Asthma (wheeze): If < 3 years with wheeze, **yellow goes to hospital** and green needs morning clinic follow-up. If **>= 3 years**, 100 ug/puff 2 puffs with **spacer** every 4 h for 5 days for wheeze.<br><br>If less than 1 year and significant mucus, provide bulb suction and saline drops. ☐ yes ☐ no | |
| 96. Dehydration Vomit #3A Diarrhea #3 (☐ not needed) | ORS Correction (75ml/kg), Amount = _ _ _ ☐ L ☐ Packets; ☐ over 4 hrs ☐ over 6 hr<br>ORS Maintenance, Amount = _ _ _ ☐ Liters ☐ Packets. [½ glass to 1 glass per loose stool or vomit; also give ORS for patients with hydration concerns – no tears/ urine]<br><br>*Diarrhea*. For cholera, antibiotic are for 'Some'/ 'Severe'. Don't delay transport for antibiotics.<br>**Watery: <8y**:Erythromycin 12.5mg/kg per dose by mouth 4 x a day for 3 days; max dose 500mg<br>**Watery: >=8 y**: Doxycycline 4 mg / kg mg by mouth once; maximum dose is 300 mg<br>**Bloody: (all ages).** Azithromycin: 10 mg/kg by mouth once a day for 5 days; max dose 500 mg<br><br>*Diarrhea present*. **Zinc:**<br>☐ 2mo-6mo. 10 mg by mouth once a day for 10 days.<br>☐ 6mo-5years. 20 mg by mouth once a day for 10 days. | |
| 97. Ear Pain #4 (☐ not needed) | First choice (ear pain <14 days). Amoxicillin: 40 mg/kg per dose by mouth twice daily for 5 days. | |
| 98. Skin #5 (☐ not needed) | Allergic (Benadryl/Hydrocortisone). Infection (Cefalexin). Scabies. (Benzyl Benzoate). | |
| 99. Pain with urine #6 (☐ not needed) | First choice. Co-trimoxazole: 10 mg/kg trimethoprim and 40 mg/kg sulfamethoxazole per dose twice a day for 5 days. If pneumonia is also present, use amoxicillin/ clavulanic acid. | |
| 100. Other problem (#7) (☐ not needed)<br>Note: Strep throat requires 10 days of Amoxicillin. | Medications (see clinical guideline appendix) | |
| 101. Additional actions (☐ not needed) | | |
| 102.Where to treat? | ☐Hospital ☐Household (yellow/green) | 103. If in 5 km, delivery needed:<br>☐ Yes, ☐ No |
| 104. Morning follow-up | ☐ Clinic (mandatory)<br>☐ Clinic if not improved (most yellow)<br>☐ Clinic as needed (most green)<br>☐ Not applicable | 105. Notes |

#### CHANGES MADE AT THE HOUSEHOLD ☐ REFUSED/ OUT OF AREA/ FAILED

| | | |
|---|---|---|
| 106. RE-ASSESSMENT | ☐ No change ☐ **A. Mild** ☐ **B. Moderate** ☐ **C. Severe** (hospital). | |
| 107. Medication change | ☐ No change Problem type (____________________) and assessment change:<br>Medication change: | |
| 108.Where to treat? | ☐ No change ☐ Hospital ☐ Household (yellow/green) | 109. Time nurse returns to the office:<br>__ __ : __ __ am ☐ pm☐ |
| 110. Morning follow-up | ☐ No change<br>☐ Clinic (mandatory)<br>☐ Clinic if not improved (most yellow)<br>☐ Clinic as needed (most green) | 111. Notes<br><br>112. GPS N____.______ W____.______ |

**Text 2.** Clinical Guidelines

These guidelines were written to be used within the scope of this pilot only

##### *A.* WEIGHT ESTIMATES

Most pediatric treatment plans require weight based dosing. There are three equations to estimate the weight:

· For 0-12 months (new APLS): weight in kg = [ age in months x 0.5 ] + 4.

· For 1-5 years (old APLS formula): weight in kg = [ age in years + 4 ] x 2.

· For 6-12 years (Luscombe and Owen Formula): weight in kg = [ age in years x 3 ] + 7.

**Decision support.** Decision support tools will be provided to the medical team. These will include the WHO IMCI guidelines, American Academy of Pediatrics, Pediatric Telephone Protocols (Schmitt, 16th Edition), pediStat (calculates medication doses), and the Outbreak Responder rehydration calculator developed by Dr. Nelson et al.

##### B. CLINICAL MANAGEMENT BY PROBLEM

**#1. Fever with or without a source.**

The World Health Organization (WHO) IMCI guidelines divide management of fever between children less than two months of age and patients older than two months of age. The reason is that young infants can be at risk of life-threatening illness and may show very subtle symptoms and signs of impending death. Therefore, escalation of care is often recommended for infants less than 2 months of age. Most call reports of fever by parents on the phone will be subjective (not measured). These subjective fevers will be considered a legitimate fever until proven otherwise.

The phrase ‘without’ source refers to patients that do not have a clear explanation for their fever (e.g. viral cold with nasal discharge, pain from urination from a bacterial urinary tract infection, skin infection, invasive diarrhea with blood in stool). This section prioritizes care for patients with fever / low temperature (hypothermia) without a source. For those patients older than 2 months of age with a source, the recommendations for that specific problem are listed in other sections. Be mindful that young children with urinary tract infections will present with fever without a source because they will not complain, or be able to complain, of pain with urination. If this is a concern, they need to be seen at a clinical for a urine test and given antibiotics in the morning.

**Less than 2 months of age.**

1. **RED. Danger sign or general clinical concern** (not feeding well, convulsions, fast breathing of 60 breaths per minute or faster), severe chest indrawing (retractions), measured fever (>=38 C rectal; >= 37.5 C oral), measured low temperature (less than 35.5 rectal or oral), movement only when stimulated (lethargic) or no movement (unresponsive). The case report form provides age-specific parameters for high and low rates; see vital sign table.

   Action: Will need immediate referral to hospital.

   Follow-up: Not applicable (will be at hospital).

2. **YELLOW / GREEN. No danger sign and no general clinical concern.**

   Action: Given that danger signs include a fever, most cases will be referred to a hospital. If there is borderline fever and mild family/ clinical concern, these cases can be monitored, supported (paracetamol and ORS) and seen at the clinic in the morning. Give 1 liter of ORS via bottle to bottle fed patients as needed to avoid dehydration; breastfed patients should continue to breastfeed. The patient should take the ORS only if they are unable to feed normally. Once made, keep for only 8 hours.

   Medication/ fluid dosing

   Paracetamol: 15 mg/ kg per dose by mouth every 6 hours as needed for fever and discomfort.

   ORS: Use as needed to avoid dehydration.

   Follow-up: Have the patient follow up with the clinic in the morning.

**Older than 2 months of age.**

1. **RED. Danger sign or stiff neck.** See above and case report form for more information.

   Action. Will need immediate referral to hospital.

   Follow up: Not applicable (will be at hospital).

2. **YELLOW / GREEN. No danger sign and no stiff neck.** See above and case report form for more information.

Action.

Give paracetamol for fever.

Give 1 liter of ORS to all patients to be used as needed to avoid dehydration. The patient should take the ORS only if they are unable to feed normally. Once made, keep for only 8 hours.

If there is a source, see that specific section for antibiotic recommendations.

Medication/ fluid dosing

Paracetamol: 15 mg/ kg per dose by mouth every 6 hours as needed for fever and discomfort.

ORS: Use as needed to avoid dehydration.

Follow-up: If there is no improvement by the morning, have the patient follow up with the clinic that morning.

**#2. Breathing problem / Cough.**

The WHO IMCI guidelines focus on identifying if the patient has respiratory distress or respiratory failure. These are assessed by looking for signs of work of breathing: fast breathing, chest indrawing showing ribs (retractions), abnormal sound when the child breaths in (e.g. stridor), wheezing when the child breaths out (e.g. asthma), nasal flaring or head bobbing. Causes of breathing problems are bacterial pneumonia, asthma attack, stridor from croup (upper airway inflammation), foreign body (object in airway), and common viral respiratory infection/ cold.

1. **RED. Danger sign, when patient is calm has abnormal sounds when breathing in (stridor), severe fast breathing (under 1 year >= 60 breaths per min; 1 year or older >=50 breaths per min) and/ or has three signs of distress (head bobbing, chest indrawing, nasal flaring). The case report form provides age-specific parameters for high and low respiration and respiration rates; see also vital sign table.**

   Action. Will need immediate referral to hospital.

   Follow up: Not applicable (will be at hospital)

2. **YELLOW. No danger sign but mild fast breathing for age and/or two signs of distress (head bobbing, chest indrawing, or nasal flaring). Upper limit for mild fast breathing: under 1 year 50 to less than 60 breaths per min; 1 year or older < 40 to less than 50 breaths per min.**

   Action

   If fever (measured either at home or by nurse) and cough, give amoxicillin.

   If 3 or more years old with wheeze or asthma history, give salbutamol.

   If less than 3 years old with wheeze or asthma history, patient goes to hospital because salbutamol needs a nebulizer.

   If less than 1 year old and significant mucus, suction the nose with a bulb and saline drops.

   Give 1 liter of ORS to all patients to be used as needed to avoid dehydration. The patient should take the ORS only if they are unable to feed normally. Once made, keep for only 8 hours.

   Provide supportive care with safe remedies for sore throat or cough.

   Medication/ fluid dosing

   Amoxicillin 40 mg/kg per dose by mouth twice daily for 5 days. Maximum single dose is 1000 mg.

   Salbutamol (100 ug/puff) two puffs with a spacer every 4 hours for 5 days.

   Paracetamol 15 mg/ kg per dose by mouth every 6 hours as needed for fever and discomfort.

   ORS: Use as needed to avoid dehydration.

   Follow-up: If there is no improvement by the morning, have the patient follow up with the clinic that morning. Patients over 3 years of age with wheeze, should follow-up with the clinic in the morning.

3. **GREEN. No danger sign.**

   Action:

   If fever (measured either at home or by nurse) and cough, give amoxicillin.

   If 3 or more years old with wheeze or asthma history, give salbutamol.

   If under 3 years old, needs morning clinic follow-up.

   If less than 1 year old and significant mucus, suction the nose with a bulb and saline drops.

   Provide supportive care with safe remedies for sore throat or cough.

Give 1 liter of ORS to all patients to be used as needed to avoid dehydration. The patient should take the ORS only if they are unable to feed normally. Once made, keep for only 8 hours.

Provide supportive care with safe remedies for sore throat or cough.

Medication/ fluid dosing

Amoxicillin 40 mg/kg per dose by mouth twice daily for 5 days. Maximum single dose is 1000 mg.

Salbutamol (100 ug/puff) two puffs with a spacer every 4 hours for 5 days.

Paracetamol 15 mg/ kg per dose by mouth every 6 hours as needed for fever and discomfort.

ORS: Use as needed to avoid dehydration..

Follow-up: Follow up if patient remains without danger signs and does not worsen. For patients under 3 years of age with wheeze, morning follow-up is required.

**#3. Dehydration/ Vomit (3A)/ Diarrhea (3B).**

The WHO IMCI guidelines address the assessment and treatment of dehydration in the context of diarrheal disease. However, methods of rehydration for diarrheal disease can also be used to address dehydration from other causes, including vomiting. Determining if a child has tears (best for children under 5 years) or has urinated (all ages) in the last 8 hours can be useful to determine risk of dehydration. With respect to vomiting, vomiting alone is not dangerous, especially if it follows a cough in a young child. Dark green vomit however can be a sign of intestinal obstruction. Multiple episodes in 24 hours (approximately above six), and/ or if child cannot take liquids, can lead to dehydration and electrolyte problems.

Assessment. The assessment of dehydration is based on four clinical features: general condition (well/alert, restless/irritable (<5 years only), lethargic/unconscious), eyes (normal/sunken), thirst (normal, drinks eagerly/ thirsty, not able to drink/ drinks poorly), skin turgor (goes back quickly in less than two seconds, slowly two to three seconds, very slowly greater than three seconds). These signs are scored ‘No’, ‘Some’, and ‘Severe’ to approximate 0-4%, 5-9%, and 10% weight loss by scoring two features in the highest category.

SEVERE (10+). At least two signs of Severe dehydration (10+%): Lethargy/unconscious, eyes very sunken, drinks poorly/ unable to drink and/or skin pinch goes back very slowly/ >3 seconds. This is RED.

SOME (5-10%). At least two signs of Some dehydration (5-10%): Restless/irritable (only for < 5 years), sunken eyes, drinks eagerly/ thirsty, and/ or skin pinch goes back slowly/ 2-3 seconds. This is YELLOW.

NO (0-5%): Default if above criteria are not met. This is GREEN.

Treatment:

1. **RED. Danger sign, severe dehydration and/or dark green vomit**.

   Action. Will need immediate referral to hospital.

   Follow up: Not applicable (will be at hospital)

2. **YELLOW. No danger sign and Some dehydration.**

   Action

   Give ORS. If patient has vomiting. Give more frequent ORS in smaller volumes.

   Give antibiotics if indicated (read below).

   Consider anti-vomiting medication if available and vomiting is severe.

   Medication/ fluid dosing

   Give ORS. The volumes for correction are as follows:

   - Volume (<1 yr): Correct with 75 ml/kg over 6 hours.
   - Volume (>=1 yr): Correct with 75 ml/kg over 4 hours.
   - Above 70 kg, the calculations are set to 70 kg (approximately 5 liters)

     While correcting for dehydration, ongoing losses must be given to replace the equivalent volume lost: Less than 2 years, 50-100 ml of ORS after each loose stool. Greater than 2 years, 100-200 ml of ORS after each loose stool.

   Give Antibiotic for patients with acute watery diarrhea that looks like rice-water and is concerning for cholera (no blood).

Below 8 years: Erythromycin 12.5 mg / kg per dose by mouth four times a day for 3 days; maximum single dose is 500 mg. Note that Doxycycline can alternatively be given to children less than 8 years per WHO guidelines.

Above 8 years: Doxycycline 4 mg / kg mg by mouth once; maximum dose is 300 mg.

Give azithromycin for patients with stool/ diarrhea with blood.

Azithromycin: 10 mg / kg by mouth once a day for 5 days; max single dose 500 mg.

Alternative: Ciprofloxacin per dose 15 mg/ kg by mouth twice a day for three days; maximum single dose is 500 mg.

Give zinc. For 2 months to 6 month old, 10 mg by mouth once a day for 10 days. For 6 months to five years, 20 mg by mouth once a day for 10 days.

Give ondansetron (if available and for severe emesis or nausea): For 1 year to 4 years, 2mg by mouth once. For 4 years and above, 4mg by mouth once.

<u>Follow-up</u>: If there is no improvement by the morning, have the patient follow up with the clinic that morning.

3. **GREEN. No danger sign and No dehydration.**

<u>Action</u>

Give ORS. If patient has vomiting, give more frequent ORS in smaller volumes. For children that don't have diarrhea but do not have tears or have not urinated in the last 8 hours, give 1-2 packets of ORS.

Give antibiotics only for patients with blood in the stool (not for acute watery diarrhea).

Consider anti-vomiting medication if available and vomiting is severe.

<u>Medication/ fluid dosing</u>

Give ORS.  Ongoing losses from diarrhea are replaced with the equivalent volume lost. Similarly, each episode of emesis can be replaced with an equal volume of ORS but space in smaller volumes over time.

- Less than 2 years, 50-100 ml of ORS after each loose stool.
- Greater than 2 years, 100-200 ml of ORS after each loose stool.

Give azithromycin for patients with stool/ diarrhea with blood.

Azithromycin: 10 mg / kg by mouth once a day for 5 days; max single dose 500 mg.

Alternative: Ciprofloxacin 15 mg/ kg per dose by mouth twice a day for three days; maximum single dose is 500 mg.

Give ondansetron (if available and for severe emesis or nausea): For 1 year to 4 years, 2mg by mouth once. For 4 years and above, 4mg by mouth once.

<u>Follow-up</u>: No need for follow up if patient remains without danger signs and does not worsen.

**<u>#4</u>. Ear pain.**

Ear pain in pediatrics is typically caused by fluid congestion in the inner ear and/or inflammation from a viral infection, and less often from a bacterial infection. It is typically not an emergency unless there is infection into the surrounding tissues. Ear pain can also be referred from pain in the mouth.

- RED. Pain when you press the bone behind the ear may indicate mastoiditis that will require hospital level care. The area may be red, warm and swollen in mastoiditis (infection of the mastoid boney space).
- YELLOW.

  Acute infection: Ear pain and/or puss draining from the ear for less than 14 days. Give amoxicillin 40 mg/kg per dose by mouth twice daily for 5 days. Treat pain with acetaminophen and /or ibuprofen.

  Chronic Infection: These should be managed at a clinic. Puss is draining for more than 14 days. Place cotton wick in the ear to drain the puss. Use topical quinolone drops for 14 days.
- GREEN. If there is no ear pain and no puss, then no treatment is needed.

<u>Follow-up</u>: Yellow follow up at clinic if no improvement. Green follow up as needed.

**#5 Skin problem.**

Skin problems may be caused by an allergy after an exposure (e.g. a specific food with a known food allergy), infection (e.g. fungus, virus, bacteria) or irritation from an insect (e.g. scabies, bed bugs). Most of these problems will not require seeking care at night. However, rapidly spreading infections or infections near the mouth, eyes, or genitalia may require attention. A bacterial infection is characterized by being red, warm, raised, and painful; not all infections are with fever; they are also often a specific location with or without puss. The size of the infection is important (coin-size, size of a hand, larger than the size of a hand). Sometimes fluid forms below the skin causing an abscess that might be infected with bacteria.

- RED. Skin infections that are spreading quickly and/or involve the eye, mouth, and genitalia (other than mild fungal diaper rash) may need care at the hospital night. Large infections (larger than the size of the hand) that might be bacterial may need hospital level care. Allergic reactions that have abdominal pain, swollen tongue, severe hives or lips require immediate loratadine and/or diphenhydramine and transfer to the hospital for concern of anaphylaxis.
- YELLOW. Skin infections that are specific to one part of the body and are red, warm, raised, and painful with fever are likely to be bacterial. Initiate treatment with Cephalexin (25 mg/kg per dose orally twice a day for five days. Alternatively, a secondary choice is co-trimoxazole (trimethoprim 4 mg/kg per dose by mouth every 12 h; dosed by trimethoprim component) for 5 days. Allergic reactions with moderate hives but no abdominal pain, swollen tongue or lips can be treated with loratadine or diphenhydramine.
- GREEN. All other cases do not necessitate night-time evaluation, and should be followed up at a daytime clinic. If a non-urgent skin problem is found at the household visit, medications available are permethrin (scabies), miconazole (fungal diaper rash), and hydrocortisone (minor inflammation). Diphenhydramine can help to address itchy skin but it lasts for only a few hours, does not solve the problem and makes patients sleepy. Allergic reactions with mild hives but no abdominal pain, swollen tongue or lips can be treated with loratadine. Lice and Scabies can be treated with permethrin or benzyl benzoate. For infants, dilute 15% benzyl benzoate one-part to one-part in water, or dilute 25% benzyl benzoate on- part to three-parts in water . Severe scabies infections with pustules can be treated with oral ivermectin only for patients that weigh at least 15 kg (0.2mg/kg per dose by mouth once; repeat in two weeks).

Follow-up: Yellow follow-up at clinic if no improvement. Green follow-up as needed.

**#6 Urinary tract infection.**

Older patients may have pain with urination (dysuria). However, young patients have more nonspecific signs. Symptoms may include fever without a source, vomiting and poor feeding, lethargy, irritability, abdominal pain, and pain on side of the lower back. Infants with fever under two-months of age will be referred to the hospital based on the fever guidelines.

- RED. Danger signs. Immediately seek care at the hospital.
- YELLOW. For pain with urination and /or lower back pain on the side. Give oral co-trimoxazole (trimethoprim 10 mg/kg by mouth every 12 h; dosed by trimethoprim component) for 5 days. Alternatively, cephalexin can be given (25 mg/ kg per dose by mouth every 6 hours for 5 days; max dose 2000 mg). If there is streptococcus pharyngitis and a urinary tract infection, consider amoxicillin/clavulinic acid 20mg/ kg by mouth every 8 hours for 10 days (dosing based on amoxicillin component).

GREEN. For pain with urination. Give oral co-trimoxazole (trimethoprim 10 mg/kg by mouth every 12 h; dosed by trimethoprim component) for 5 days. Alternatively, cephalexin can be given (25 mg/ kg per dose by mouth every 6 hours for 5 days; max dose 2000 mg). If there is streptococcus pharyngitis and a urinary tract infection, consider amoxicillin/clavulinic acid 20mg/ kg by mouth every 8 hours for 10 days (dosing based on amoxicillin component). Follow-up in clinic as needed.

Follow-up: Yellow follow-up at clinic if no improvement. Green follow-up as needed.

**E. Other diagnosis.** The management of other less common diagnoses will not be scripted. The approach will rely on standard of care by nurses, scope of practice for nurses, and consult from the on-call doctor. The intentionally short list of available medications will also constrain the scope of management to pediatric urgent care needs. One example is an allergic reaction. Using the same triage approach (RED, YELLOW, GREEN), triage the situation. If there is allergic reaction with involvement of the mouth or ability to breath, the patient is RED and needs to go to the hospital immediately; dose with diphenhydramine and loratadine during transport. If there is a full body allergic reaction without involvement of the mouth or breathing the patient is YELLOW and use diphenhydramine and loratadine can both be used. Follow with clinic in the morning if no improvement. Be aware these medications will make the patient tired. If there is an allergic reaction on a specific body part without involvement of the mouth or breathing the patient is GREEN and diphenhydramine or loratadine can both be used. Be aware diphenhydramine will make the patient tired. A second example is Strep throat; symptoms are typically fever, swollen lymph nodes along the side of the neck, no cough, and white painful lesions on the back of the throat. The dose of amoxicillin is the same (40mg/kg per dose twice a day) but the course is 10 days. For each of these situations use a similar method for follow-up: Yellow follow up at clinic if no improvement. Green follow up as needed.

**Appendix**

**Normal Vital signs***

| Age | 1-7 days | 1-3 weeks | 1 month | 6 months | 12 months | 18 months |
|---|---|---|---|---|---|---|
| Weight | 3 kg | 3 kg | 4 kg | 7 kg | 10 kg | 11 kg |
| Heart Rate | 90-165 | 105-180 | 120-180 | 100-180 | 100-180 | 100-180 |
| Resp. Rate | 30-60 | 30-60 | 30-60 | 30-60 | 24-40 | 24-40 |

| Age | 2 years | 3 years | 5 years | 6 years | 8 years | 10 years |
|---|---|---|---|---|---|---|
| Weight | 12 kg | 14 kg | 18 kg | 25 kg | 31 kg | 37 kg |
| Heart Rate | 60-140 | 60-140 | 60-140 | 60-140 | 60-140 | 60-140 |
| Resp. Rate | 24-40 | 24-40 | 22-34 | 18-30 | 18-30 | 18-30 |

*Fever: oral >37.5 C; rectal >38 C. Oxygen: Normal >=90%. Normal values derived from PALS and Stanford (LPCH) Pediatric Emergency Guidelines. Weight estimates explained at the top of the guidelines. WHO IMCI thresholds set fast breathing at 50 breaths per minute (<1 year) and 40 breaths per minute (>=1 year).

### MotoMeds Formulary (updated 5/20/2020; for both under and over 10 years)

| Name | Formulation | Indication | Potential common side-effect | Dosage |
|---|---|---|---|---|
| **1 Non-opioids and non-steroidal anti-inflammatory medicines (NSAIMs)** | | | | |
| Ibuprofen | **Oral liquid:** 200 mg/5 mL<br>**Tablet:** 200 mg; 400 mg; 600 mg | Inflamation/ pain/ fever | Stomach irritation | Children 10 mg/kg per dose by mouth every 6 hours as needed.<br>Adults: 400 mg by mouth every 6 hours as needed. |
| Paracetamol | **Oral liquid:** 125 mg/5 mL<br>**Tablet:** 100 mg to 500 mg<br>**Suppository:** 100 mg | Fever/ pain/ inflamation | No common side effect | Children: 15 mg/kg per dose by mouth every 6 hours as needed.<br>Adults: 500 mg by mouth every 6 hours as needed. |
| **2. ANTIALLERGICS AND MEDICINES USED IN ANAPHYLAXIS** | | | | |
| Loratadine | **Oral liquid:** 1 mg/mL<br>**Tablet:** 10 mg | Mild allergy/ allergic reaction | Mild tiredness | 2-5 years old: 5 mg by mouth once daily.<br>5-18 years old: 10 mg by mouth once daily.<br>Adults: 20 mg by mouth once daily. |
| Diphenhydramine HCl | **Oral liquid:** 12.5mg/5mL<br>**Tablet:** 25 mg | Mild allergy/ allergic reaction | Moderate tiredness | 2-5 years old: 6.25 mg by mouth every 6 hours as needed for allergic reaction.<br>5-10 years old: 12.5 mg by mouth every 6 hours as needed for allergic reaction.<br>Over 10 years: 25 mg by mouth every 6 hours as needed for allergic reaction. |
| **3. Anti-bacterials** | | | | |
| ***3.1 Beta-lactam medicines*** | | | | |
| Amoxicillin | **Powder for oral liquid:** 125 mg (as trihydrate)/5 mL; 250 mg (as trihydrate)/5 mL | Ear, lung, systemic infection | Rash/ allergic reaction | Children: 40 mg/kg per dose by mouth twice daily for 5 days.<br>Adults: 875 mg by mouth every 12 hours for 5 days. |
| Amoxicillin / clavulanic acid | **Powder for oral liquid:** ___ mg Amox / ____ Clavulanic Acid per 5 mL | Lung infection when amoxicllin fails or is not available, or there is a UTI plus lung infection. | Rash/ allergic reaction | Children: 20 mg/ kg every 8 hours for 10 days. Dose based on amoxicillin component.<br>Adults: 250 mg by mouth every 8 hours for 5 days. |
| Azithromycin | **Tablet:** 500 mg | Bloody diarrhea, watery diarrhea concerning for cholera, respiratory infection. | Rash/ allergic reaction | **For bloody diarrhea:** children 10 mg / kg by mouth once a day for 5 days (max 500 mg); adults 500 mg by mouth once a day for 5 days.<br>**For watery diarrhea**: chilldren 20 mg / kg by mouth as a single dose; adults (>15 yrs or >50kg) 1000 mg by mouth as a single dose.<br>**Bacterial pneumonia** (primary or secondary): children 10 mg/kg (max 500mg) per dose by mouth once, then 5 mg/kg (max 250mg) by mouth daily for days 2-5; adults 500mg by mouth once, then 250 mg by mouth daily for days 2-5. |
| Cefalexin (Cephalexin) | **Powder for reconstitution with water:** 125 mg/5 mL; 250 mg/5 mL (anhydrous).<br>**Solid oral dosage form:** 250 mg (as monohydrate). | Skin infection, urinary tract infection | Rash/ allergic reaction | **Skin Infection:** children 25 mg/kg per dose by mouth twice a day for five days; adults adults 500 mg every 12 hours for 5 days.<br>**Urinary tract infection:** children 25 mg/kg per dose by mouth four times a day for 5 days; adults 500 mg twice a day for 5 days. |
| ***3.2 Other antibacterials*** | | | | |
| Ciprofloxacin | **Tablet:** 500 mg | Bloody diarrhea, urinary tract infection | Rash/ allergic reaction. Tendonitis. | **For bloody diarrhea:** children 15 mg/ kg by mouth twice a day for three days; adults 500 mg by mouth twice a day for three days. |
| Doxycycline | **Tablet:** ___ mg | Infection, including watery diarrhea concerning for cholera | Rash/ allergic reaction | **For watery diarrhea (cholera):** chilren 4 mg/ kg mg by mouth once; adults 300 mg by mouth once. |
| Erythromycin | **Powder for oral liquid:** 125 mg/5 mL (as stearate **or** estolate **or** ethyl succinate)<br>**Solid oral dosage form:** 250 mg (as stearate **or** estolate **or** ethyl succinate) | Infection, including watery diarrhea concerning for cholera<br>Infection, including watery diarrhea concerning for cholera | Rash/ allergic reaction<br>Rash/ allergic reaction | **For watery diarrhea (cholera) for children less than 8 years**: Erythromycin 12.5 mg / kg per dose by mouth four times a day for 3 days; maximum single dose is 500 mg |
| Levofloxacin | **Tablet:** ___ mg | Moderate to severe rimary / secondary bacterial pneumonia (COVID associated). | Rash/ allergic reaction | **For watery diarrhea (cholera):** chilren 4 mg/ kg mg by mouth once; adults 300 mg by mouth once. |
| sulfamethoxazole + trimethoprim | **Oral liquid:** 200 mg + 40 mg/5 mL.<br>**Tablet:** 100 mg + 20 mg; 400 mg + 80 mg | Infection, including skin and urinary tract infection (UTI). | Rash/ allergic reaction. Avoid in children less than 2 months. | **UTI:** children 10 mg/kg trimethoprim per dose by mouth twice a day for 5 days (WHO); adults sulfamethoxazole 800 mg/trimethoprim 160 mg orally twice daily for 3 days.<br>**Skin infection/ abscess**: children 4 mg/kg trimethoprim per dose by mouth twice a day for 5 days; adults sulfamethoxazole 800 mg/trimethoprim 160 mg orally twice daily for 5 days.(Doses based on the trimethoprim component) |
| **4. Anti-fungal medicines** | | | | |
| Nystatin (oral) | **Lozenge:** 100 000 IU<br>**Oral Liquid:** 50 mg/5mL; 100000 IU/mL | Oral thrush (fungus) | Rash/ allergic reaction | Thrush: 2 ml by mouth four times a day until resolution |
| **5. DERMATOLOGICAL MEDICINES (topical)** | | | | |
| **6.1 Anti-infective medicines** | | | | |
| Antibiotic cream (neomycin sulfate, bacitracin zinc and polymyxin B) | **Cream** | Bacterial Skin infection | Rash/ allergic reaction | Apply topically as needed 3-4 times daily |
| Miconazole (topical) | **Cream or ointment:** 2% (nitrate) | Fungal skin infection (diaper rash) | Rash/ allergic reaction | Apply topically as needed 3-4 times daily |
| **6.2 Anti-inflammatory and anti-pruritic medicines** | | | | |
| Hydrocortisone (topical) | **Cream or ointment:** 1% (acetate) | Mild inflammation in skin. | None | Apply topically as needed 3-4 times daily |
| **6.3 Scabicides and pediculicides** | | | | |
| Benzyl benzoate (topical) | **Lotion:** 25% | Insect irritation (lice, scabies) | Rash/ allergic reaction | **Scabies:** Apply cream from hairline to toes once at bedtime. Rinse in morning. Repeat in 1 week.<br>**Pediculosis (lice):** Apply cream to hair at bedtime. Rinse in the morning. Repeat in 1 week.<br><br>Wash bedding and clothes with hot water. |
| Ivermectin | **Oral liquid: ___ mg /5 ml**<br>**Table: ___ mg tablet** | Scabies (severe) | Rash/ allergic reaction | Children: 0.2 mg /kg per dose by mouth once. Repeat in two weeks. Only for children that weigh at least 15 kg. Take with food. |
| Permethrin (topical) | **Cream:** 5%<br>**Lotion:** 1% | Insect irritation (lice, scabies) | Rash/ allergic reaction | **Scabies**: Apply 5% cream from hairline to toes once at bedtime. Rinse in morning. Repeat in 1 week.<br>**Pediculosis (lice):** Apply 1% cream to hair. Rinse after 1 hour. Repeat in 1 week.<br><br>Wash bedding and clothes with hot water. |
| **7. Fluids and medicines used in diarrhoea** | | | | |
| ***7.1 Oral rehydration*** | | | | |
| Oral rehydration solution | **Powder for dilution** in 200 mL; 500 mL; 1 L | Rehydration/ Hydration | None | See guidelines |
| ***7.2 Medicines for diarrhoea*** | | | | |
| Zinc sulfate | **Solid oral dosage form:** 20 mg | Shortens diarrhea course | None | 2 mo - 6 mo: 10 mg by mouth once daily for 10 days.<br>6 mo - 5 years: 20 mg by mouth once daily for 10 days. |
| **8.1 Anti-asthmatic medicines** | | | | |
| Salbutamol | **Injection:** 50 micrograms (as sulfate)/mL in 5-mL ampuole.<br>**Metered dose inhaler (aerosol):** 100 micrograms (as sulfate) per dose<br>**Respirator solution for use in nebulizers:** 5 mg (as sulfate)/mL | Asthma/ reactive airway disease. | Increases heart rate. Tremor. | 3 years and older: Two puffs (100 ug/puff) with a spacer every 4 hours for 5 days for wheeze. Under three years with yellow sign and wheeze need hospital referral; green sign and wheeze need morning clinic follow-up. |
| **9.1 Anti-emetic** | | | | |
| Ondansetron | **Oral dissolvable tab (ODT):** 4mg or 8 mg<br>**Oral liquid:** 4 mg / 5 ml | Nause | None | 1-4 years: 2 mg by mouth once<br>>=4 years: 4 mg by mouth once |
